# Dirichlet process mixture models to estimate outcomes for individuals with missing predictor data: application to predict optimal type 2 diabetes therapy in electronic health record data

**DOI:** 10.1101/2022.07.26.22278066

**Authors:** Pedro Cardoso, John M. Dennis, Jack Bowden, Beverley M. Shields, Trevelyan J. McKinley, the MASTERMIND Consortium

**Affiliations:** University of Exeter, Medical School

**Keywords:** Dirichlet process mixture model, treatment selection model, precision medicine, Type 2 diabetes, Bayesian modelling

## Abstract

**Background:** Missing data is a common problem in regression modelling. Much of the literature focuses on handling missing outcome variables, but there are also challenges when dealing with missing predictor information, particularly when trying to build prediction models for use in practice.

**Methods:** We develop a flexible Bayesian approach for handling missing predictor information in regression models. For prediction this provides practitioners with full posterior predictive distributions for both the missing predictor information and the outcome variable, conditional on the observed predictors. We apply our approach to a previously proposed treatment selection model for type 2 diabetes second-line therapies. Our approach combines a regression model and a Dirichlet process mixture model (DPMM), where the former defines the treatment selection model and the latter provides a flexible way to model the joint distribution of the predictors.

**Results:** We show that under missing-completely-at-random (MCAR) and missing-at-random (MAR) assumptions (with respect to the missing predictors), the DPMM can model complex relationships between predictor variables, and predict missing values conditionally on existing information. We also demonstrate that in the presence of multiple missing predictors, the DPMM model can be used to explore which variable(s), if collected, could provide the most additional information about the likely outcome.

**Conclusions:** Our approach can provide practitioners with supplementary information to aid treatment selection decisions in the presence of missing data, and can be readily extended to other types of response model.

**Key Messages:**

- Missing predictor variables present a significant challenge when building and implementing prediction models in clinical practice.
- Removing individuals with missing information and performing a complete case analysis can lead to imprecision and bias. Multiple imputation approaches typically translate uncertainty through prediction model parameter standard errors, as opposed to a consistent joint probability model.
- Alternatively, a Bayesian approach using Dirichlet process mixture models (DPMMs) offers a flexible way to model complex joint distributions of predictor variables, which can be used to estimate posterior (predictive) distributions for the missing predictors, conditional on the observed predictors.
- Using a DPMM, in this way allows uncertainties around missing predictor data to be propagated through to a prediction model of interest using a Bayesian hierarchical framework. This allows prediction models to be developed using datasets with incomplete predictor information (assuming missing-completely-at-random/missing-at-random). Furthermore, predictions can be made on new individuals even if they have incomplete predictor information (under the same assumptions).
- This approach provides full posterior predictive probability distributions for both missing predictor variables and the outcome variable, allowing a wide range of probabilistic models outputs to be derived to support clinical decision making.

## 1 Introduction

Prediction models are becoming more widely developed and used as clinical decision aids. Handling missing predictor variables is a significant challenge when building and implementing prediction models in clinical practice. Due to difficulties modelling missing data, many prediction models use only complete case information leading to biases when the data is not missing completely at random (MCAR) [1]. The use of only complete cases reduces the utility of models in clinical practice where information on certain variables is often missing for certain patients. A common approach for dealing with missing data, under the assumption of missing at random (MAR), involves the use of multiple imputation, commonly through chained equations (MICE) [2]. Whilst powerful and straightforward to implement, MICE has some limitations. Unless the chained equations have a carefully chosen hierarchical dependence structure, these do not always provide a probabilistically consistent joint probability model for the predictors [3]. Another limitation of multiple imputation involves the propagation of uncertainties to the parameters and predictions, as this only occurs through standard errors and therefore does not provide the richness-of-information afforded by full posterior distributions. These limitations have been listed as key priority areas for research in the Predictive Approaches to Treatment effect Heterogeneity (PATH) statement: to “Better understand the effect of different missingness mechanisms, and develop principled methods for dealing with missing data in the context of subgroup identification” and “Determine methods to permit models predicting treatment effect to cope with missing data in clinical practice” [4].

Hierarchical Bayesian approaches offer advantages over standard frequentist methods currently in use. The implementation of a hierarchical Bayesian prediction model, in which the response model is dependent on a flexible joint probability model for the predictor variables, facilitates both inference and prediction in the presence of missing data. Dirichlet process mixture model (DPMM) [5] can be used to capture the joint distribution of the predictor variables. This flexible model can readily capture non-standard relationships such as non-linearity and heteroscedasticity across different dimensions. Furthermore, it can handle mixtures of both numerical and categorical predictors [6, 7]. This allows for fully probabilistic inference under MCAR and MAR; addressing two key points of further research from the PATH statement. Although DPMMs have been established as a clustering mechanism [8] and used in multiple imputation [9, 10, 11], its use in regression modelling [7] is more limited. A key approach is that of *profile regression*, where a DPMM is fitted to a set of predictor variables, which generate clusters of mixture components, and the cluster indexes are then regressed against a response variable of interest [6].

In this paper we demonstrate a novel implementation of a hierarchical Bayesian prediction model using a DPMM to model the predictor variables and the predictor’s missingness. We obtain probabilistically consistent posterior distributions for the parameters and posterior predictive distributions for the missing predictors and response variables. Our approach differs from previous approaches in that we use the DPMM to generate conditional predictor information, given the observed data, which then propagates through to any model that relies on such information. We show how this approach can be used for both improved modelling of missing predictor information and better exploration of outcomes. We use an example of a treatment selection model in type 2 diabetes (T2D), which combines routine clinical features of each patient to predict future blood glucose (HbA1c) levels under two commonly prescribed treatments, SGLT2i and DPP4i [12]. The analysis identified marked heterogeneity in HbA1c outcome that is predicted by baseline patient characteristics: age-at-treatment, baseline HbA1c, body mass index (BMI), estimated glomerular filtration rate (eGFR), alanine aminotransferase (ALT) measurements and previous treatment history. These predictions can determine which treatment is likely to be optimal for an individual patient. A limitation of the current SGLT2i–DPP4i treatment selection model is the requirement for complete predictor information to make predictions for an individual, and our approach aims to address this challenge. Furthermore, the model can be used to identify how predictions of the response or treatment selection outcomes might change if any of the missing predictor variable(s) were available, and if so, which ones (if any) would be the most useful to collect. We note that this latter feature could be particularly useful for modelling non-routinely collected diagnostic test information, such that one could examine how predictions might change if the test were to be conducted, and hence could be used to inform whether it is worthwhile to perform the test or not.

## 2 Methods

### 2.1 Study Overview

This paper’s focus is on enhancing the previously described penalised linear regression model designed to predict likely glucose-lowering response (HbA1c) in people with T2D initiating SGLT2i or DPP4i therapy [12]. For an individual patient, the difference in the predicted outcome for each treatment is then used to infer which is likely to be optimal. Such models are labelled treatment selection models [13].

We note that the models developed in [13, 12] are not causal; instead they aim to model how HbA1c levels change in patients undergoing each of the two treatments separately. The model uses interaction effects between the treatment and the other predictors to allow for differential response to treatment given a set of predictors. For a new individual with a specific set of characteristics, mean predictions of the HbA1c change at 6/12 months post treatment can be made under both treatments, and then the difference can be derived. We note that there is considerable residual uncertainty in the fitted models, and as such [13, 12] only use point predictions when making comparisons. As such, predictions should be interpreted as the expected difference for an average individual with a specific set of predictors under the two treatments. In the Bayesian model below, we follow the same principle, where posterior predictive distributions are for an *average* individual with a given set of characteristics (so we do not include the individual-level residual uncertainty in these distributions).

In the model description below, we use **Y** to denote the outcome variable, and **X** to denote the set of predictor variables. Hence, *π*(**Y** | **X, *ψ***) corresponds to the likelihood function from the regression model of Dennis *et al*. [12], with parameters ***ψ***; *π*(**X** | **Θ**) corresponds to the joint probability density function for the DPMM, with parameters **Θ** (see Box 1); and *π*(***ψ*, Θ**) corresponds to the prior distribution for the parameters, then, for complete data, our Bayesian hierarchical model has posterior distribution:

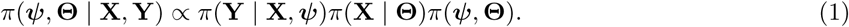

We fit this model using Markov chain Monte Carlo (MCMC) with the package ***Nimble*** [14, 15] (version 0.12.2) in the software ***R*** [16] (version 4.0.3). Code for the Bayesian model can be found in the GitHub repository https://github.com/PM-Cardoso/DPMM-tsm. We cannot share the original data due to confidentiality constraints, but we provide an example of using the code to fit a model to a smaller synthetic dataset in the Supplementary Materials.

#### Box 1.

**Dirichlet process mixture models (DPMMs)**

A DPMM is commonly used as a prior distribution over the components of a (possibly multivariate) mixture model of unknown complexity [17]. A DPMM consists of a theoretically infinite number of components, where each component is parameterised by a specific functional form with component-specific parameters. Instead of fitting an infinite number of components, we use a truncated DPMM with a maximum number of components *K*, based on the assumption the optimal number of components is lower than the set limit [18]. In turn, the computational demands for fitting the model are reduced.

The DPMM thus defines a weighted sum of *K* component densities [19]. The component densities are restricted to a particular parametric class of densities that are assumed to be appropriate for the data at hand. We define *π*_*k*_ (*X* | **Θ**_*k*_) as the *k*^th^ component density, with **Θ**_*k*_ representing the component parameters. A *K* component mixture density is defined as:

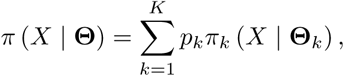

where *p*_*k*_ are component-specific weights such that 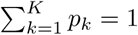 [20].

For *J*_*C*_ continuous predictors, we use mixtures of multivariate Gaussian distributions with *J*_*C*_ dimensions, the cluster specific parameters for component *k* (*k* = 1, …, *K*) are given by (***µ***_*k*_, **Σ**_*k*_), where ***µ***_*k*_ is a *J*_*C*_-vector of means and **Σ**_*k*_ is a (*J*_*C*_ × *J*_*C*_) covariance matrix. For *J*_*D*_ categorical predictors, we use mixtures of categorical probability mass functions, where the number of categories for a covariate *j* (*j* = 1, …, *J*_*D*_) is *K*_*j*_, the component-specific parameters are the probabilities of belonging to each category, given by ***ϕ***_*k*_ = (***ϕ***_*k*1_, ***ϕ***_*k*2_, …, ***ϕ***_*k*_*J*_*D*_) with ***ϕ***_*kj*_ = (*ϕ*_*kj*1_, *ϕ*_*kj*2_, …, *ϕ*_*kj*_*K*_*j*_) and 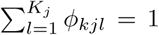. The model in this paper is given as a mixture of continuous and categorical variables, since 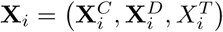. Hence in the notation of Section 2.1 **Θ** = (**Θ**_1_, …, **Θ**_*K*_), where the component-specific parameters are given by **Θ**_*k*_ = (***µ***_*k*_, **Σ**_*k*_, ***ϕ***_*k*_).

A latent variable, *Z*_*i*_ = 1, …, *K*, is used to assign individual data points to different components of the mixture model, and we assume independence between continuous and categorical components conditional on the cluster allocations [6, 7]. Thus the probability density for individual *i*, given *Z*_*i*_ is:

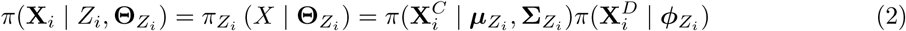

More details on the component densities, prior distributions, and how to sample from the DPMM are given in the Appendix.

### 2.2 Treatment selection model

The model from Dennis *et al*. [12] assumes a continuous, normally distributed response variable *Y*_*i*_ for *i* = 1, …, *N*, where *N* corresponds to the number of observations. The model is a standard linear regression model with both continuous and categorical predictors (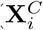 and 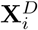 respectively). Restricted cubic splines are placed on the continuous predictors 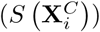 [21], and interaction terms are included between the binary drug taken 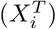 and the other variables. Hence the structure of the treatment selection model is:

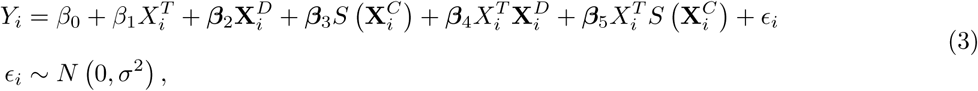

where *β*_0_ is the intercept, ***β*** = (*β*_1_, ***β***_2_, …, ***β***_5_) is a vector of regression coefficients (where ***β***_2_, …, ***β***_5_ are themselves vectors of regression coefficients corresponding to the different discrete, spline and interactions variables as required), with *σ* the residual standard deviation. Hence, in the notation of Section 2.1, **Y** = (*Y*_1_, …, *Y*_*n*_), **X** = (**X**_1_, …, **X**_*n*_), with 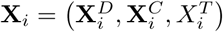 and ***ψ*** = (*β*_0_, ***β***, *σ*).

### 2.3 Data/Study Population

The analysis in this paper is performed with anonymised UK primary care electronic healthcare records from Clinical Practice Research Datalink (CPRD) GOLD (July 2019 download) [22], selecting users of SGLT2i and DPP4i therapy after January 1st, 2013. We applied the same criteria to extract clinical features (age-at-treatment, baseline HbA1c, BMI, eGFR, ALT, the number of previously prescribed glucose-lowering therapies, the current therapy and its duration, the number of ongoing prescribed treatments) and HbA1c outcome as previously described by Dennis *et al*. in the initial model development study. The dataset is split into two groups: the development group (60% of the dataset corresponding to 16126 patients) and the validation group (40% of the dataset corresponding to 10751 patients). There are an extra 2057 patients in the development group and 1375 patients in the validation group with incomplete predictor variables (Table 1) compared to Dennis *et al*. model [12]. We fit to the development dataset, and undertake both internal and external validation.

**Table 1.**
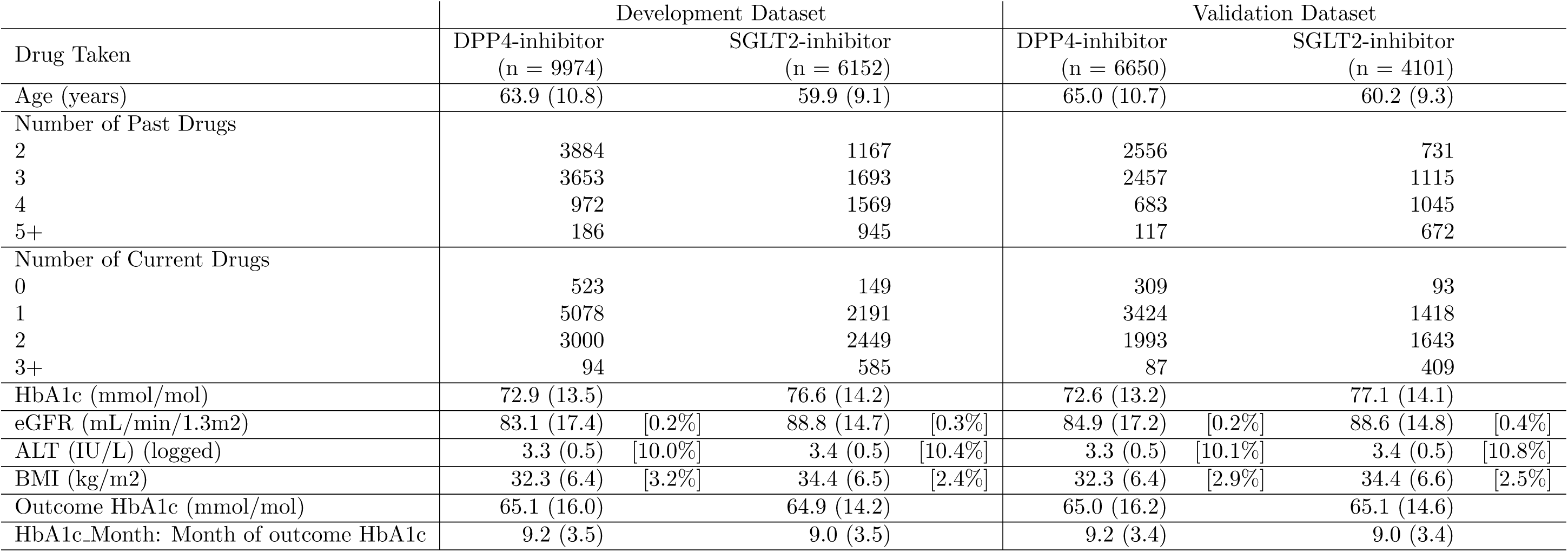
Summaries of covariates and biomarkers for the development and validation datasets. (SD) [percentage of missing values]

### 2.4 Model Validation

To assess convergence and mixing of the MCMC chains we perform a visual inspection of the trace plots for all parameters, as well as monitoring the Gelman-Rubin 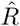 values [23]—see Figures S2–S3. The Gelman-Rubin 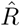values for *α, σ* and regression parameters vary between 1 and 1.005. These suggest that the chains converged and are mixing well.

Since the application of DPMM in the model uses a truncated number of clusters, we also inspect the posterior mean number of individuals in components ranked by occupancy (Figure S2), alongside a posterior predictive plot of the fitted DPMM against the empirical distribution of the predictors from the development and validation data sets (Figures S4 and 1 respectively). Contrasting standard prediction models, the principal aspect of a treatment selection model is to accurately predict the optimal treatment instead of necessarily accurately predicting the outcome [24].

Validation of the individualised treatment effects of the model is carried out according to the guidelines provided by Dennis *et al*. [12]. The first method suggested is verifying the prediction performance by checking the slope of predicted HbA1c versus the observed outcome—Figure S6. This shows that there is no systematic bias in the mean function, but that there is considerable residual variation as per the results of [12]. The second method is to plot a density graph for all patients regarding the difference between each therapy’s treatment effects (Figure S7), that can be compared to [12]. This shows the predicted number of patients benefiting from DPP4i or SGLT2i across the population and shows good agreement with [12]. The final method involves defining two subgroups of patients: a concordant subgroup whose treatment was the same as their predicted optimal treatment and a discordant subgroup whose treatment did not coincide with their predicted optimal treatment, and then plotting their treatment response across time (Figure S8), which demonstrates that the concordant group experience a higher therapy benefit than the discordant group.

## 3 Results

### 3.1 Fit and validation of the DPMM

To visualise the fit of the DPMM, we take the bivariate marginal posterior predictive distributions for a combination of variables versus the empirical equivalent (Figure 1). The combination of density plots, density map plots and bar plots demonstrate that the fitted DPMM provides a good representation of both the development (Figure S4) and external validation (Figure 1) datasets.

**Figure 1:**
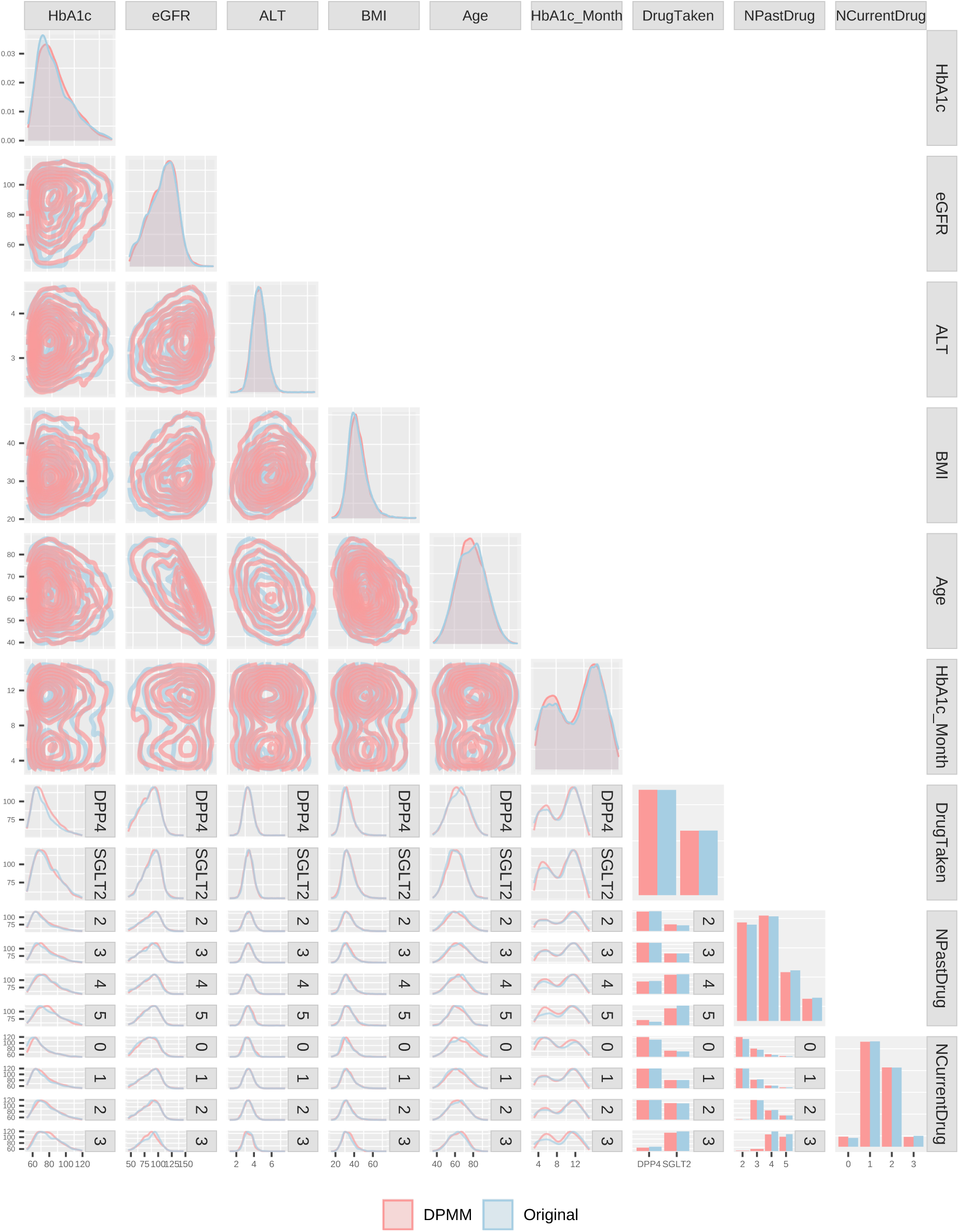
Generalised pairs plot of predictor variables for an independent validation dataset against an equal number of random samples from Dirichlet process mixture model (DPMM). The DPMM provides a exceptional representation of the validation dataset.

### 3.2 Probabilistic predictions using the Bayesian treatment selection model

The Bayesian model produces full posterior predictive distributions that marginalise the uncertainty in the parameters and any missing predictor variables. This means we can make probabilistic statements about the parameter values and predicted outcomes, based on the observed data and the choice of prior distributions, in contrast to frequentist models where uncertainty statements relate to the expected long-term behaviour of an estimator if the same study were to be repeated *ad infinitum* [25]. In Figure 2, we showcase the probability distributions of average therapy response for three anonymised patients (A, B and C). The model predicts a 98% probability of DPP4i performing better than SGLT2i on average for patient A. For patient B, SGLT2i has a 42% probability of performing better on average than DPP4i. If the patient’s characteristics are similar to patient C, the model predicts SGLT2i as the optimal treatment on average with a 99% probability. An alternative way to visualise this information is by utilising probability distributions for treatment response difference (Figure 3). With this approach, it is evident that there is a high probability of patient A having a 1–3 mmol/mol greater HbA1c reduction on average with SGLT2i instead of DPP4i. Patient B is predicted a possible average HbA1c benefit between 0–3 mmol/mol for both therapies. For patient C, there is a high probability of DPP4i resulting in a greater HbA1c reduction than SGLT2i, with a likely 1–7 mmol/mol benefit on average. As discussed earlier, these are posterior predictive distributions for the *mean* response for a patient with each set of characteristics (see Box 2).

**Figure 2:**
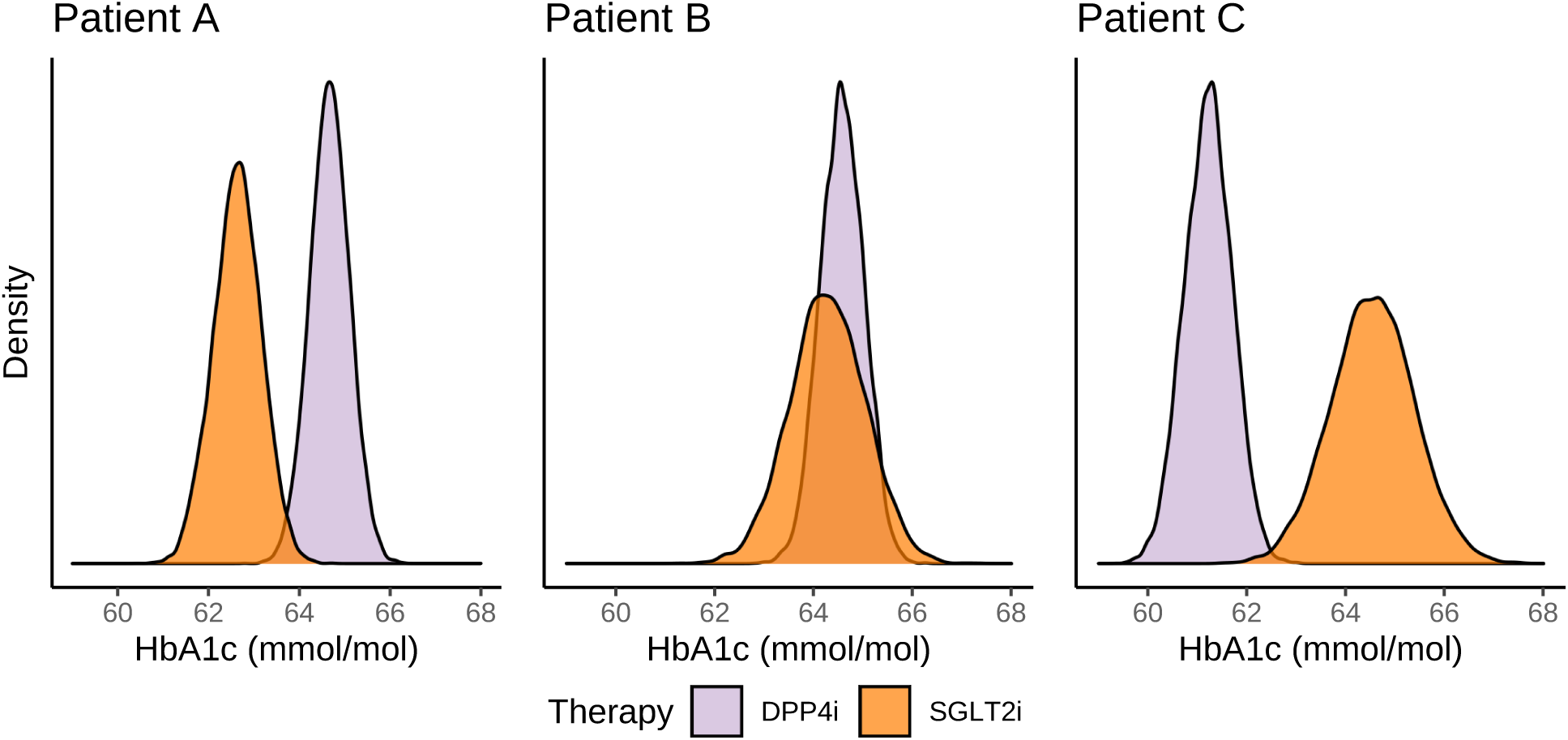
Therapy probability distribution at 6 month prediction. The model is capable of providing a range of treatment responses depending on the patient’s data. For patient A, DPP4i has a 98% probability of performing better than SGLT2i. For patient B, SGLT2i has a 42% probability of performing better than DPP4i. For patient C, SGLT2i has a >99% probability of performing better than DPP4i. Patient [A,B,C]: Number of Past Drugs [4,3,4], Number of Current Drugs [2,2,2], HbA1c [67,75,65], eGFR [84.2,66.6,67.9], ALT(log) [3.4,2.8,2.6], BMI [26.1,33.4,28.5], Age [68,79,81]

**Figure 3:**
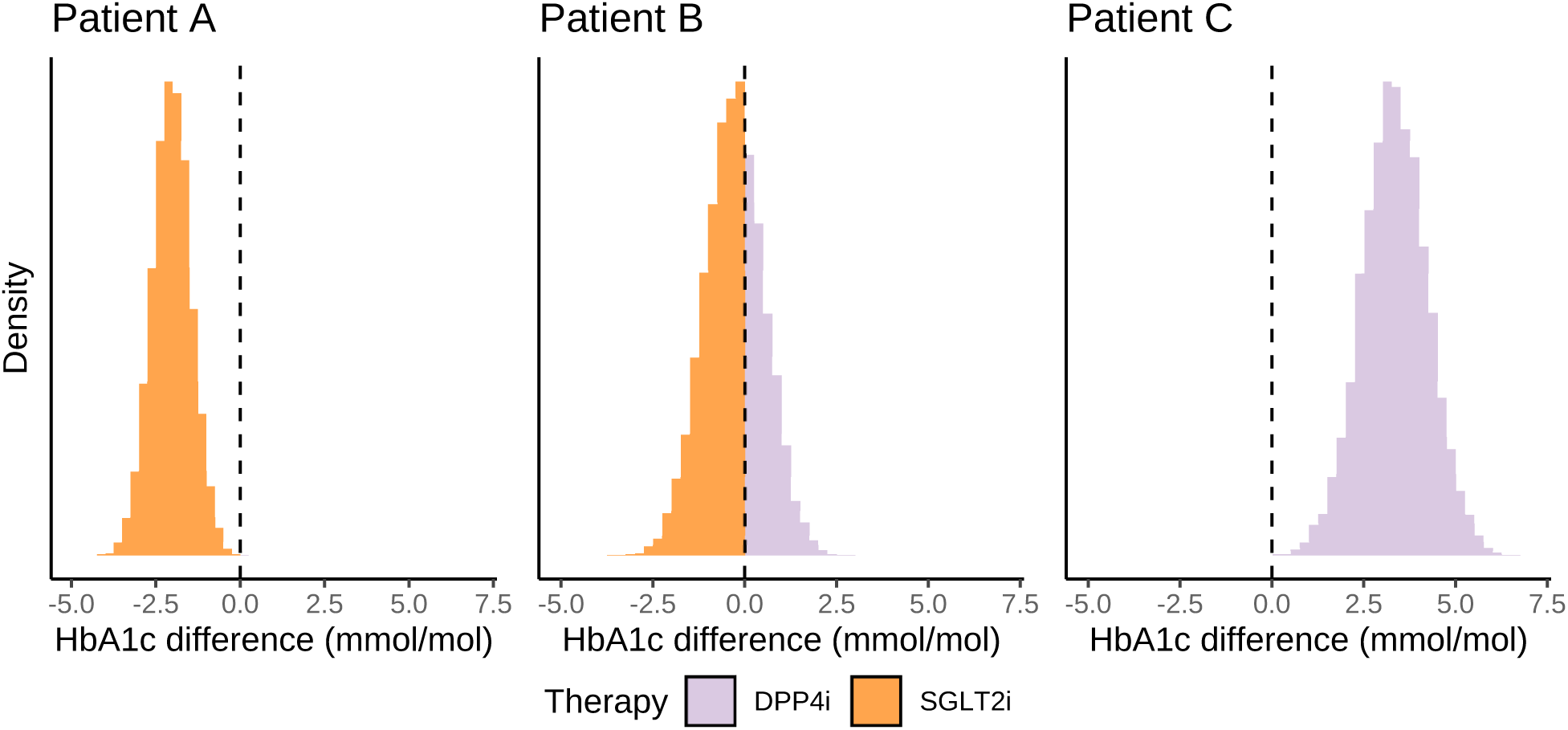
Predicted treatment response difference at 6 months. A negative value corresponds to a benefit on SGLT2i and a positive value corresponds to a benefit on DPP4i. Patient [A,B,C]: Number of Past Drugs [4,3,4], Number of Current Drugs [2,2,2], HbA1c [67,75,65], eGFR [84.2,66.6,67.9], ALT(log) [3.4,2.8,2.6], BMI [26.1,33.4,28.5], Age [68,79,81]

#### Box 2.

**Posterior predictive distributions**

In Bayesian modelling it is possible to derive a full *posterior predictive distribution* for any new individual. Thus the uncertainty associated with a prediction is captured through a probability distribution, from which point estimates can be derived, or alternatively probabilistic questions can be asked, such as those described in Section 3.2. The definition of a posterior predictive distribution for an individual with complete predictor information is:

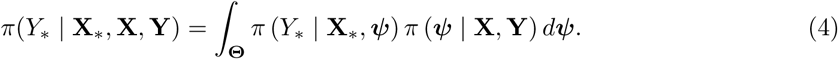

Thus the predictive distribution *integrates* (or *averages*) over the posterior distribution for the parameters, and thus naturally incorporates the uncertainties about the parameters as well as those arising from the underlying model. The examples in this paper present the posterior predictive distribution for an average individual, with a given set of characteristics, defined by:

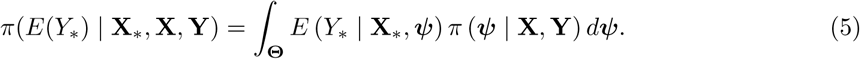

Equations (4-5) are analogous to a confidence interval and point prediction for the model response. We are free to use either approach or both, dependent on the context. Please see the Appendix for more information about how different predictive distributions can be derived and sampled from (including when predictors are missing).

### 3.3 Predictions for patients with missing data using the Bayesian treatment selection model

The inclusion of DPMM in the Bayesian model enables it to make predictions for patients with missing information. These predictions are possible by averaging over the posterior distributions for the missing values. The posterior distributions are conditional on the existing data, and the uncertainty of the distributions is in part related to the degree of missingness. Figure 4 demonstrates how the data available for patient A influences the uncertainty of the posterior predictive distributions for the missing variables and the response. For each scenario, different sets of predictor variables are artificially missing. Each row displays how the uncertainty of the posterior predictive distributions for the missing covariates change as more data becomes available. The uncertainty of posterior predictive treatment response distribution decreases as more variables become available. As baseline HbA1c is the most influential predictor in the model, the availability of the baseline HbA1c value vastly reduces the uncertainty of the prediction compared to other variables.

**Figure 4:**
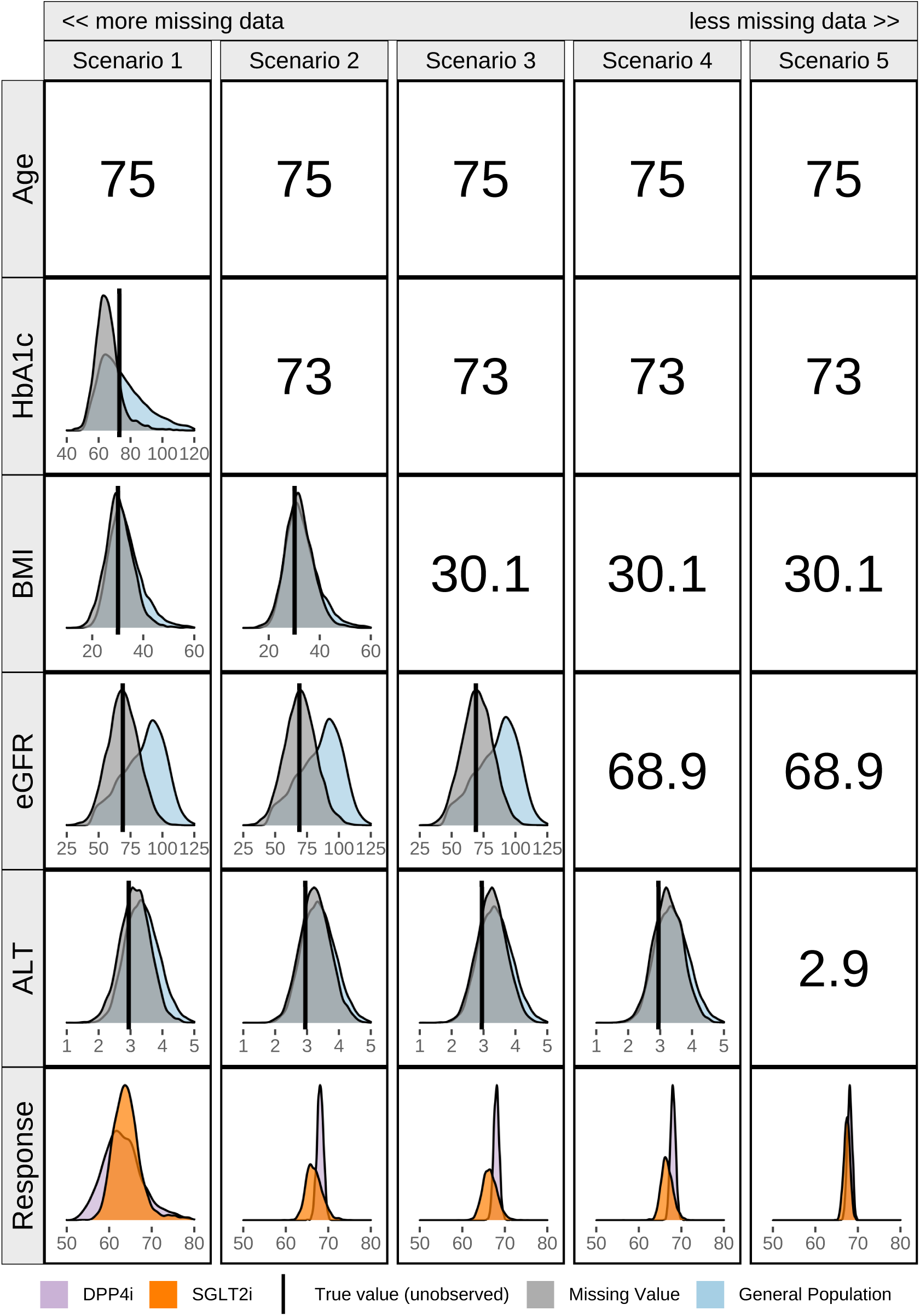
Predictive distributions for missing data and treatment response conditional on the scenario for a patient at 6 months. Unobserved true values are given by the black vertical line, and its prediction by the grey distribution. As more variables become known, the uncertainty in the predictions decreases. Scenario 1: Past Drugs, Current Drugs, Age known. Scenario 2: Past Drugs, Current Drugs, Age, HbA1c known. Scenario 3: Past Drugs, Current Drugs, Age, HbA1c, BMI known. Scenario 4: Past Drugs, Current Drugs, Age, HbA1c, BMI, eGFR known. Scenario 5: All variables known. Patient: Number of Past Drugs [2], Number of Current Drugs [0], HbA1c [73], eGFR [68.9], ALT(log) [2.9], BMI [30.1], Age [75]

### 3.4 Variable influence on the response prediction using the Bayesian treatment selection model

Alternatively, due to the model providing posterior predictive distributions for missing data, a new question can be proposed—which missing variable will, if collected, give the most information about the treatment response? Here is a case where it is necessary to know which variable information to gather next. In this example, the patient only has three predictors available (number of previous therapies, current therapies and age), as all other predictor variables are missing. With three predictors, the model predicts similar treatment responses for both DPP4i and SGLT2i. For each missing predictor, a range of treatment response distributions can be estimated for different values of the predictors, chosen as different quantiles from the corresponding conditional posterior predictive distributions given the observed predictors. This provides an insight into how the treatment response prediction might be affected by a range of predictor values.

Figure 5 showcases the conditional probability distributions for any missing variable, and we can explore what we might expect to see at different values informed by these distributions. For example, when taking low, medium and high quantile values for HbA1c, it is evident that the treatment response drastically changes. For this patient, if HbA1c takes the 5% quantile value, there is a high probability the treatment response is below 65 mmol/mol, with DPP4i therapy expected to perform better than SGLT2i therapy. On the other hand, if HbA1c takes the 50% quantile value, the expected HbA1c response is around 60–80 mmol/mol and with no distinction between the treatment response for both therapies. Lastly, if HbA1c takes the 95% quantile value, the expected treatment response is likely above 70 mmol/mol, without differentiation between both treatments. In contrast, when other missing variables take different quantile values, it results in very similar treatment responses across all quantiles. All of the above provides evidence that HbA1c will be the most useful additional variable to collect to improve the choice of therapy for this patient.

**Figure 5:**
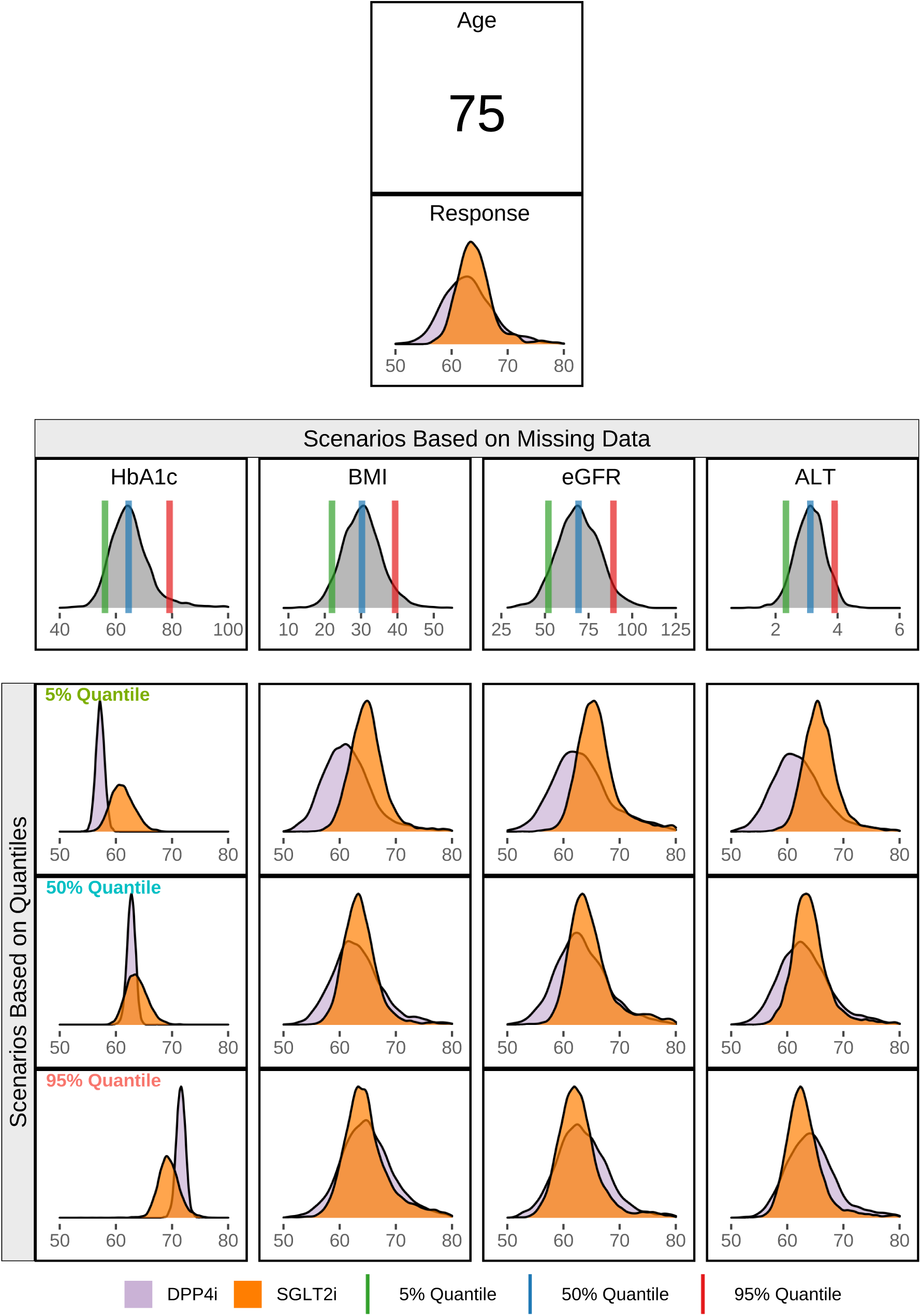
Analysis of variable influence on response prediction for a patient at 6 months. The initial response prediction is conditional on age, number of previous and current therapies. Further predictions are conditional on quantile values for HbA1c, BMI, eGFR and ALT. Out of the 4 missing covariates, collection HbA1c will, in theory, provide the most information about treatment response for the patient. Patient: Number of Past Drugs [2], Number of Current Drugs [0], Age [75]

### 3.5 High-risk patient prediction using the Bayesian treatment selection model

In addition to all previous aspects, the Bayesian model could also be used in a different way. For example, in the case of a high-risk patient, it may be desirable to choose the treatment response that is most likely to result in a reduction of HbA1c to below 60 mmol/mol. Hence we could choose a treatment on the basis of overall predicted benefit, or we could select the therapy with the least probability of exceeding the target threshold. Using Figure S9 as an arbitrary example, there is a high probability of SGLT2i resulting in a better treatment response than DPP4i for this patient. In a conventional approach, SGLT2i would be the therapy chosen. However, it is critical to note that SGLT2i has a higher probability of resulting in a value of HbA1c above 60 mmol/mol than DPP4i. In this instance, the fact that DPP4i is less likely to result in a value above the target could be more influential in the choice of therapy than SGLT2i being more likely to result in lower treatment response on average. Choosing DPP4i as a therapy for this patient gives the highest probability of an average treatment response below the target.

## 4 Discussion

This paper provides details on how to fit a Bayesian treatment selection model with continuous and categorical covariates using a Dirichlet process mixture model. Crucially, the model enables model fitting and response prediction for patients with complete and incomplete data (under the assumption values are MCAR/MAR). Furthermore, it can be used to gain insight into the utility of collecting further data on patients with missing predictor values.

The Bayesian treatment selection model presented in this paper augments the classical model developed by Dennis et al. [12]. The model validation is carried out the same way as the original version. This validation assesses whether the treatment selection model accurately predicts the optimal treatment rather than the exact therapy response. We use posterior mean predictions in an analogous way to the classical point prediction used in [12]. With that in mind, the performance of the Bayesian model is comparable. The regression parameter values are similar between the three models, with the Bayesian model achieving similar regularisation through the use of weakly informative regularising priors. Including individuals with incomplete data in the model fitting process reduces the uncertainty of regression parameters marginally (Figure S5).

There are several applications for this type of approach. We have presented average patient predictions due to the high residual uncertainty of Dennis *et al*. [12], but this may not be true for all prediction models. One is calculating the treatment response alongside the difference in treatment response for both therapies. Furthermore, due to the inclusion of DPMM, the model can provide probability distributions for any missing covariates. The availability of probability distributions for missing covariates, in turn, can be used to influence the choice of future covariate data collection to improve treatment response prediction. Instead of having a set rule, we use Figure 5 to choose a variable that reduces the uncertainty in the posterior distribution for the difference between the two treatments. Alternatively, we use Figure 5 to decide on the variable that provides the highest posterior probability of preference for one therapy. Lastly, if Figure 5 shows that one variable pushes the treatment response towards a side of the scale and a second variable pulls the treatment response towards another side, then collecting both variables would be beneficial.

The application of the DPMM in the context of treatment selection illustrates multiple potential benefits of the approach when developing clinical prediction tools. It enables the ability to build prediction models in the presence of any missing data, under the assumption the data is MCAR/MAR. Furthermore, even in the presence of complete data, the inclusion of the DPMM in the model building enables the potential to make predictions for patients with incomplete predictor information. Work is underway to expand the model when the missingness mechanism is not MAR/MCAR. The extension of the model allows for simultaneous fitting of the missingness mechanism, the treatment selection model and the DPMM. This is a novel approach with considerable advantages for prediction from treatment selection models, and many of these advantages would also be true for standard clinical prediction models.

## Supporting information

Sample Code

## Data Availability

All data produced in the present study are available upon reasonable request to the authors.

## 6 Acknowledgements

This article is based in part on data from the Clinical Practice Research Datalink (CPRD) obtained under licence from the UK Medicines and Healthcare products Regulatory Agency. CPRD data is provided by patients and collected by the NHS as part of their care and support.

## 7 Data Availability Statement

The data underlying this article cannot be shared publicly due to the privacy of individuals that participated in the study. Synthetic sample data is available of GitHub within the repository “PM-Cardoso/DPMM-tsm”.

## 8 Supplementary Material

Supplementary data are available online.

## References

[1] S.X. Lee J.G. McLachlan, S. Rathnayake. Comprehensive chemometrics: chemical and biochemical data analysis. Elsevier, 2 edition, 2020.

[2] D.B. Rubin. Multiple imputation for nonresponse in surveys. New York: John Wiley & Sons, Inc, 1987.

[3] J.A. Melissa, A.S. Elizabeth, F. Constantine, and J.L. Philip. Multiple imputation by chained equations: what is it and how does it work? Int J Methods Psychiatr Res, 20(1):40–49, 2011.

[4] D.M. Kent, J.K. Paulus, D. van Klaveren, and et al. The predictive approaches to treatment effect heterogeneity (PATH) statement. Ann Intern Med, 172(35), 2020.

[5] J.D. McAuliffe, D.M. Blei, and M.I. Jordan. Nonparametric empirical bayes for the dirichlet process mixture model. Stat Comput, 16:5–14, 2006.

[6] J. Molitor, M. Papathomas, M. Jerrett, and S. Richardson. Bayesian profile regression with an application to the national survey of children’s health. Biostatistics, 11(3):484–498, 2010.

[7] S. Liverani, D.I. Hastie, L. Azizi, M. Papathomas, and S. Richardson. Premium: An R package for profile regression mixture models using dirichlet processes. J Stat Softw, 64(7):1–30, 2015.

[8] Y. Li, E. Schofield, and M. Gönen. A tutorial on dirichlet process mixture modeling. J Math Psychol, 91:128–144, 2019.

[9] M. Di Zio, U. Guarnera, and O. Luzi. Imputation through finite gaussian mixture models. Comput Stat Data Anal, 51(11):5305–5316, 2007.

[10] H.J. Kim, J.P. Reiter, Q. Wang, L.H. Cox, and A.F. Kar. Multiple imputation of missing or faulty values under linear constraints. J Bus Econ Stat, 31(2):375–386, 2014.

[11] Y. Si and J.P. Reiter. Nonparametric bayesian multiple imputation for incomplete categorical variables in large-scale assessment surveys. J Educ Behav Stat, 38(5):499–521, 2013.

[12] J. Dennis, K. Young, A. Mcgovern, and et al. Derivation and validation of a type 2 diabetes treatment selection algorithm for sglt2-inhibitor and dpp4-inhibitor therapies based on glucose-lowering efficacy: cohort study using trial and routine clinical data. medRxiv. preprint: not peer reviewed.

[13] J. Dennis. Precision medicine in type 2 diabetes: using individualized prediction models to optimise selection of treatment. Diabetes, 69:2075–2085, 2020.

[14] P. de Valpine, D. Turek, C. Paciorek, and et al. Programming with models: writing statistical algorithms for general model structures with NIMBLE. J Comput Graph Stat, 26:403–413, 2017.

[15] P. de Valpine, C. Paciorek, D. Turek, and et al. NIMBLE: MCMC, particle filtering, and programmable hierarchical modeling, 2022. R package version 0.12.2.

[16] R Core Team. R: a language and environment for statistical computing. R Foundation for Statistical Computing, Vienna, Austria, 2021.

[17] T.S. Ferguson. A bayesian analysis of some nonparametric problems. Ann Stat, 1(2):209–230, 1973.

[18] S. Favaro and S.G. Walker. A generalized constructive definition for the dirichlet process. Stat Probab Lett, 78(16), 2010.

[19] G. McLachlan and D. Peel. Finite mixture models. New York: John Wiley Sons, 1 edition, 2000.

[20] O. Papaspiliopoulos and G.O. Roberts. Retrospective markov chain monte carlo methods for dirichlet process hierarchical models. Biometrika, 95(1):169–186, 2008.

[21] F.E. Harrell Jr. Regression modeling strategies. New York: Springer International Publishing, 2 edition, 2015.

[22] E. Herrett, A.M. Gallagher, K. Bhaskaran, and et al. Data resource profile: clinical practice research datalink (CPRD). Int J Epidemiol, 44(3):827–36, 2015.

[23] A. Gelman and D.B. Rubin. Inference from iterative simulation using multiple sequences. Stat Sci, 7(4):457–511, 1992.

[24] P.M. Bossuyt and T. Parvin. Evaluating biomarkers for guiding treatment decisions. Ejifcc, 26(1):63–70, 2015.

[25] A. Gelman, J.B. Carlin, H.S. Stern, D.B. Dunson, A. Vehtari, and D.B. Rubin. Bayesian data analysis. New York: Chapman Hall/CRC, 3 edition, 2013.

