## Supplementary Material for "Dirichlet process mixture models to estimate outcomes for individuals with missing predictor data: application to predict optimal type 2 diabetes therapy in electronic health record data"

### 1 Appendix

#### 1.1 Component distributions

As described in Box 1, for $J_C$ continuous predictors, we use mixtures of multivariate Gaussian distributions with $J_C$ dimensions, with cluster specific parameters for component $k$ ($k = 1, \ldots, K$) given by $(\boldsymbol{\mu}_k, \boldsymbol{\Sigma}_k)$, where $\boldsymbol{\mu}_k$ is a $J_C$-vector of means and $\boldsymbol{\Sigma}_k$ is a $(J_C \times J_C)$ covariance matrix. Hence

$$\pi\left(\mathbf{X}_i^C \mid Z_i, \boldsymbol{\Theta}_{Z_i}\right) = \frac{1}{\sqrt{(2\pi)^J \mid \boldsymbol{\Sigma}_{Z_i} \mid}} \exp\left[-\frac{1}{2}\left(\mathbf{X}_i^C - \boldsymbol{\mu}_{Z_i}\right)^T \boldsymbol{\Sigma}_{Z_i}^{-1}\left(\mathbf{X}_i^C - \boldsymbol{\mu}_{Z_i}\right)\right]. \tag{1}$$

For $J_D$ categorical predictors, we use mixtures of categorical probability mass functions, where the number of categories for a covariate $j$ ($j = 1, \ldots, J_D$) is $K_j$, the component-specific parameters are the probabilities of belonging to each category, given by $\boldsymbol{\phi}_k = (\boldsymbol{\phi}_{k1}, \boldsymbol{\phi}_{k2}, ..., \boldsymbol{\phi}_{kJ_D})$ with $\boldsymbol{\phi}_{kj} = (\phi_{kj1}, \phi_{kj2}, \ldots, \phi_{kjK_j})$ and $\sum_{l=1}^{K_j} \phi_{kjl} = 1$. Hence

$$\pi(\mathbf{X}_i^D \mid Z_i, \boldsymbol{\Theta}_{Z_i}) = \prod_{j=1}^{J} \boldsymbol{\phi}_{Z_i j}^{X_{ij}}. \tag{2}$$

#### 1.2 Prior distributions

We use a stick-breaking [**?**, **?**] construction for the prior probabilities of the mixture components $p_k$. Conceptually, this involves repeatedly breaking off and discarding a random fraction of a stick with an initial length of 1. The fraction discarded is sampled from a Beta distribution with shape parameter $\alpha$ (where $\alpha$ influences the prior weights on the number of components).

$$p_k = V_k \prod_{l<k}(1 - V_l)$$

$$p_1 = V_1$$

$$V_k \sim \text{Beta}(1, \alpha)$$

In order to improve convergence and mixing of the MCMC algorithm, all continuous variables are standardised. The regression parameters ($\boldsymbol{\beta}$) are fitted with weakly informative shrinkage priors, which acts as a regularisation technique and can reduce posterior uncertainty alongside stabilising computations [?]. The prior distributions for regression parameters and $\sigma$ are:

$$\boldsymbol{\beta} \sim \text{Normal}(0, 2.5)$$

$$\sigma \sim \text{Exponential}(1)$$

The clustering and components of the DPMM follows a hierarchical structure, with the following prior distributions:

$$\begin{aligned}
\mathbf{Z} &\sim \text{Dirichlet}(\alpha) \\
\alpha &\sim \text{Gamma}(\text{shape} = 2, \text{rate} = 1) \\
\boldsymbol{\mu}_k &\sim \text{Multivariate Normal}(\boldsymbol{\mu}_0, \boldsymbol{\Sigma}_0) \\
\boldsymbol{\Sigma}_k &\sim \text{Wishart}(\text{R}, \rho) \\
R &\sim \text{Wishart}(\text{R}_0, \rho_0) \\
\rho &\sim \text{Exponential}(0.1)
\end{aligned}$$

In order to fit the model, we set some initial values. For $\boldsymbol{\mu}_k$, we set $\boldsymbol{\mu}_0$ to be a vector with the means of continuous variables and we set $\boldsymbol{\Sigma}_0$ to be the covariance matrix corresponding to a diagonal matrix with the range of values for the continuous variables. For $R$, we set the degrees of freedom to the number of continuous variables in the model, $\rho_0 = 6$.

### 1.3 Predictive distributions

If there are no missing predictor variables, then the DPMM provides no additional information about the treatment selection model parameters, and could be integrated out of (**??**). The purpose of including the DPMM is two-fold: a) to allow incomplete predictor data to be included when the model is fitted; and b) to allow predictions to be made for individuals with missing predictor information. Hence, if we have a combination of missing ($\mathbf{X}^m$) and non-missing ($\mathbf{X}^o$) predictors, then the target posterior distribution is thus:

$$\pi(\boldsymbol{\psi}, \boldsymbol{\Theta}, \mathbf{X}^m \mid \mathbf{X}^o, \mathbf{Y}) \propto \pi(\mathbf{Y} \mid \mathbf{X}^o, \mathbf{X}^m, \boldsymbol{\psi})\pi(\mathbf{X}^o, \mathbf{X}^m \mid \boldsymbol{\Theta})\pi(\boldsymbol{\psi}, \boldsymbol{\Theta}), \tag{3}$$

and as such we get full posterior predictive distributions for all of the missing variables $\mathbf{X}^m$. We can then integrate over the missing variables to get the marginal posterior distribution for the parameters-of-interest:

$$\pi(\boldsymbol{\psi}, \boldsymbol{\Theta} \mid \mathbf{X}^o, \mathbf{Y}) = \int_{\mathbf{X}^m} \pi(\boldsymbol{\psi}, \boldsymbol{\Theta}, \mathbf{X}^m \mid \mathbf{X}, \mathbf{Y}) d\mathbf{X}^m. \tag{4}$$

This (multi-dimensional) integral can be done numerically using the posterior samples generated from the MCMC. In this sense the Bayesian model naturally propagates the uncertainties from the missing information, through to the posterior distributions for the parameters.

Similarly, since the DPMM provides a flexible joint probability model for the predictor variables, we can leverage this to produce posterior predictive distributions for a new individual with observed predictors $\mathbf{X}_*^o$ say. In this case

$$\pi(Y_*, \mathbf{X}_*^m \mid \mathbf{X}_*^o, \mathbf{X}^o, \mathbf{Y}) = \int_{\boldsymbol{\Theta}} \int_{\boldsymbol{\psi}} \pi\left(Y_* \mid \mathbf{X}_*^m, \mathbf{X}_*^o, \boldsymbol{\psi}\right) \pi\left(\mathbf{X}_*^m \mid \mathbf{X}_*^o, \boldsymbol{\Theta}\right) \pi\left(\boldsymbol{\psi}, \boldsymbol{\Theta} \mid \mathbf{X}^o, \mathbf{Y}\right) d\boldsymbol{\psi} d\boldsymbol{\Theta}, \tag{5}$$

gives the joint posterior predictive distribution for $Y_*$ and $\mathbf{X}_*^m$, where $\pi\left(\mathbf{X}_*^m \mid \mathbf{X}_*^o, \boldsymbol{\Theta}\right)$ is the conditional distribution for $\mathbf{X}_*^m$ given $\mathbf{X}_*^o$, which can be derived directly from the DPMM (see Section 1.4 for details). Again, these distributions can be estimated via Monte Carlo simulation, using the posterior samples generated from the MCMC, and then simulating from the conditional DPMM and the treatment selection model for each set of samples. Estimates of the marginal posterior predictive distributions for $\pi(Y_* \mid \mathbf{X}_*^o, \mathbf{X}^o, \mathbf{Y}, \boldsymbol{\psi})$ and $\pi(X_*^m \mid \mathbf{X}_*^o, \mathbf{X}^o, \mathbf{Y}, \boldsymbol{\Theta})$ can be readily generated in a similar way.

In the case where a new individual has complete covariate information, then the posterior predictive distribution for $Y_*$ reduces to

$$\pi(Y_* \mid \mathbf{X}_*, \mathbf{X}^o, \mathbf{Y}) = \int_{\boldsymbol{\Theta}} \pi\left(Y_* \mid \mathbf{X}_*, \boldsymbol{\psi}\right) \pi\left(\boldsymbol{\psi} \mid \mathbf{X}^o, \mathbf{Y}\right) d\boldsymbol{\psi}, \tag{6}$$

(hence has no dependence on the DPMM). The plots presented in this paper ignore the residual variation and hence use point predictions. See Box 2 for more information. Full details are given in Section 1.4 and example code for fitting the model, and generating posterior predictive samples is given in the Supplementary Material.

### 1.4 Prediction equations for conditional predictive draws

Posterior predictive distributions for patients with missing information can be generated empirically using posterior samples generated from the MCMC, as long as we can sample from a conditional DPMM and the treatment selection model (both described in Section 1.3). Suppose $\mathbf{X}_*$ is a $J$-dimensional vector of covariates for a new individual. We partition $\mathbf{X}_*$ into two disjoint subsets $\mathbf{X}_*^m$ and $\mathbf{X}_*^o$, where $\mathbf{X}_*^m$ are the missing covariates of $M$ dimensions and $\mathbf{X}_*^o$ are the observed covariates of $J-M$ dimensions. We will estimate the joint posterior predictive distribution for the response $\mathbf{Y}_*$ and $\mathbf{X}_*^m$ given the observations $\mathbf{X}_*^o$:

$$\pi\left(\mathbf{Y}_*, \mathbf{X}_*^m \mid \mathbf{X}_*^o, \mathbf{X}_{1:N}, \mathbf{Y}_{1:N}\right) = \int_{\boldsymbol{\psi}} \int_{\boldsymbol{\Theta}} \pi\left(\mathbf{Y}_* \mid \mathbf{X}_*^m, \mathbf{X}_*^o, \boldsymbol{\psi}\right) \pi\left(\mathbf{X}_*^m \mid \mathbf{X}_*^o, \boldsymbol{\Theta}\right) \pi\left(\boldsymbol{\psi}, \boldsymbol{\Theta} \mid \mathbf{Y}_{1:N}, \mathbf{X}_{1:N}^o\right) d\boldsymbol{\psi} d\boldsymbol{\Theta}, \quad (7)$$

where $\pi\left(\boldsymbol{\psi}, \boldsymbol{\Theta} \mid \mathbf{Y}_{1:N}, \mathbf{X}_{1:N}^o\right)$ is the marginal posterior distribution given the observed data $\mathbf{Y}_{1:N}$ and $\mathbf{X}_{1:N}^o$.

We can estimate (7) through Monte Carlo sampling, by first drawing $L$ random samples, $(\boldsymbol{\psi}_l, \boldsymbol{\Theta}_l)$ $(l = 1, \ldots, L)$, from the posterior distribution (obtained through the original MCMC runs), and then for each of these we sample from $\pi\left(\mathbf{X}_{*l}^m \mid \mathbf{X}_*^o, \boldsymbol{\Theta}_l\right)$ and then $\pi\left(\mathbf{Y}_{*l} \mid \mathbf{X}_{*l}^m, \mathbf{X}_*^o, \boldsymbol{\psi}_l\right)$ as detailed below.

For a given set of parameters $\boldsymbol{\Theta}$, we need to generate random samples from:

$$\pi\left(\mathbf{X}_*^m \mid \mathbf{X}_*^o, \boldsymbol{\Theta}\right) = \sum_{z=1}^{K} \pi\left(\mathbf{X}_*^m \mid Z = z, \mathbf{X}_*^o, \boldsymbol{\Theta}\right) P\left(Z = z \mid \mathbf{X}_*^o, \boldsymbol{\Theta}\right). \quad (8)$$

We can sample from (8) by first drawing a component $Z$ from:

$$\begin{aligned} P\left(Z = z \mid \mathbf{X}_*^o, \boldsymbol{\Theta}\right) &\propto \pi\left(\mathbf{X}_*^o \mid Z = z, \boldsymbol{\Theta}\right) \pi\left(Z = z \mid \boldsymbol{\Theta}\right) \qquad \text{from Bayes' Theorem} \\ &\propto \pi\left(\mathbf{X}_*^{Co} \mid \boldsymbol{\mu}_z, \boldsymbol{\Sigma}_z\right) \pi\left(\mathbf{X}_*^{Do} \mid \boldsymbol{\phi}_z\right) \pi\left(Z = z \mid \boldsymbol{\pi}, \alpha\right) \qquad \text{from (??)}, \end{aligned}$$

where $\mathbf{X}_*^{Co}$ and $\mathbf{X}_*^{Do}$ are the observed continuous and categorical variables respectively. Then, given $Z = z$, we have

$$\begin{aligned} \pi\left(\mathbf{X}_*^m \mid Z = z, \mathbf{X}_*^o, \boldsymbol{\Theta}\right) &= \pi\left(\mathbf{X}_*^{Cm} \mid Z = z, \mathbf{X}_*^o, \boldsymbol{\Theta}\right) \pi\left(\mathbf{X}_*^{Dm} \mid Z = z, \mathbf{X}_*^o, \boldsymbol{\Theta}\right) \\ &= \pi\left(\mathbf{X}_*^{Cm} \mid \mathbf{X}_*^{Co}, \boldsymbol{\mu}_z, \boldsymbol{\Sigma}_z\right) \pi\left(\mathbf{X}_*^{Dm} \mid \mathbf{X}_*^{Do}, \boldsymbol{\phi}_z\right) \quad \text{from (??)}. \end{aligned} \quad (9)$$

We can sample from (9) by taking independent random samples from $\left(\mathbf{X}_*^{Cm} \mid \mathbf{X}_*^{Co}, \boldsymbol{\mu}_z, \boldsymbol{\Sigma}_z\right)$ and $\left(\mathbf{X}_*^{Dm} \mid \mathbf{X}_*^{Do}, \boldsymbol{\phi}_z\right)$, where

$$\left(\mathbf{X}_*^{Cm} \mid \mathbf{X}_*^{Co}, \boldsymbol{\mu}_z, \boldsymbol{\Sigma}_z\right) \sim N\left(\boldsymbol{\mu}_{*z}^{m|o}, \boldsymbol{\Sigma}_z^{m|o}\right),$$

with

$$\boldsymbol{\mu}_{*z}^{m|o} = \boldsymbol{\mu}_z^m + \boldsymbol{\Sigma}_z^{mo}\left(\boldsymbol{\Sigma}_z^{oo}\right)^{-1}\left(\mathbf{X}_*^{Co} - \boldsymbol{\mu}_z^o\right)$$

$$\boldsymbol{\Sigma}_z^{m|o} = \boldsymbol{\Sigma}_z^{mm} - \boldsymbol{\Sigma}_z^{mo}\left(\boldsymbol{\Sigma}_z^{oo}\right)^{-1}\boldsymbol{\Sigma}_z^{om},$$

assuming that $\boldsymbol{\mu}_z^C$ and $\boldsymbol{\Sigma}_z^C$ are partitioned such that

$$\boldsymbol{\mu}_z^C = \begin{pmatrix} \boldsymbol{\mu}_z^m \\ \boldsymbol{\mu}_z^o \end{pmatrix} \quad \text{and} \quad \boldsymbol{\Sigma}_z^C = \begin{pmatrix} \boldsymbol{\Sigma}_z^{mm} & \boldsymbol{\Sigma}_z^{mo} \\ \boldsymbol{\Sigma}_z^{om} & \boldsymbol{\Sigma}_z^{oo} \end{pmatrix}$$

Finally, we sample

$$\left(\mathbf{X}_{*j}^{Dm} \mid \boldsymbol{\phi}_z\right) \sim \text{Multinomial}\left(\boldsymbol{\phi}_{kj}\right)$$

independently for $j = 1, \ldots, M$ missing covariates (since $\mathbf{X}_{*j}^{Dm}$ are conditionally independent of $\mathbf{X}_{*j}^{Do}$ given cluster $z$).

Once we have $L$ random samples for $\mathbf{X}_{*l}^m$, we can then sample (with a slight abuse of notation) from the treatment selection model such that $\mathbf{Y}_{*l} \sim \pi\left(\mathbf{Y} \mid \mathbf{X}_{*l}^m, \mathbf{X}_*^o, \boldsymbol{\psi}_l\right)$.

### 1.5 Convergence diagnostics

The Bayesian model was run for 50 000 iterations in two separate chains. Inspection of $\alpha$ values (Figure S2C) as well as $\sigma$ and regression parameters (Figure S3) revealed that the first twenty thousand iterations should be discarded as burn-in. The trace plots for the regression parameters demonstrate the remaining iterations after the burn-in suggest convergence of the model (Figure S3). During the model fit, not all components had patients assigned to them. After removing burn-in and ranking components by occupancy (Figure S2B), only 18 components were utilised with components below the 14$^{\text{th}}$ rank having less than 30 patients (0.2%) assigned to them (Figure S2A). The Gelman-Rubin $\hat{R}$ values for $\alpha$, $\sigma$ and regression parameters vary between 1 and 1.005.

### 1.6 The Bayesian treatment selection model is consistent with the original penalised maximum likelihood regression model

The posterior distributions for the regression parameters in the model fitted to incomplete data stay consistent with the equivalent model fitted to complete data in both Bayesian and frequentist approaches (Figure S5). The posterior credible intervals for the Bayesian model fitted to the complete data are slightly narrower on the whole than the frequentist confidence intervals, due to the weak shrinkage priors used. We can see that the inclusion of the incomplete data results in a reduction of the posterior intervals, due to an increase in the number of data points available to inform the model fit.

Internal validation of the model shows the final model explained 29.4% of the variation of HbA1c outcome, with a good calibration (slope = 1.0015 [1 = perfect]) (Figure S6A). Validation of the model in the hold-out dataset shows that the model explained 29.4% of the variation of the HbA1c outcome, alongside a good calibration (slope = 1.0200) (Figure S6B). In the development dataset, 13368 patients are predicted to benefit from SGLT2i therapy, and 2758 patients are predicted to benefit from DPP4i therapy. In cases where SGLT2i is predicted as the optimal therapy, 176 patients are predicted a benefit >10 mmol/mol and 6355 patients are predicted to benefit between 5–10 mmol/mol. Whereas when DPP4i is predicted as the optimal therapy, 316 patients are predicted a benefit >5 mmol/mol (Figure S7A). In the validation dataset, 8929 patients are predicted to benefit from SGLT2i therapy, and 1822 patients are predicted to benefit from DPP4i therapy. In cases where SGLT2i is predicted as the optimal therapy, 123 patients are predicted a benefit > 10 mmol/mol and 4198 patients are predicted to benefit between 5–10 mmol/mol. Compared with when DPP4i is predicted as the optimal therapy, 209 patients are predicted a benefit >5 mmol/mol (Figure S7B). Furthermore, Figure S8A/B demonstrates the concordant subgroup of patients has a higher therapy response than the discordant subgroup for the development and validation datasets, in both DPP4i and SGLT2i therapies. These are consistent with the results in Dennis *et al.* [**?**].

### 2 Supplementary Figures

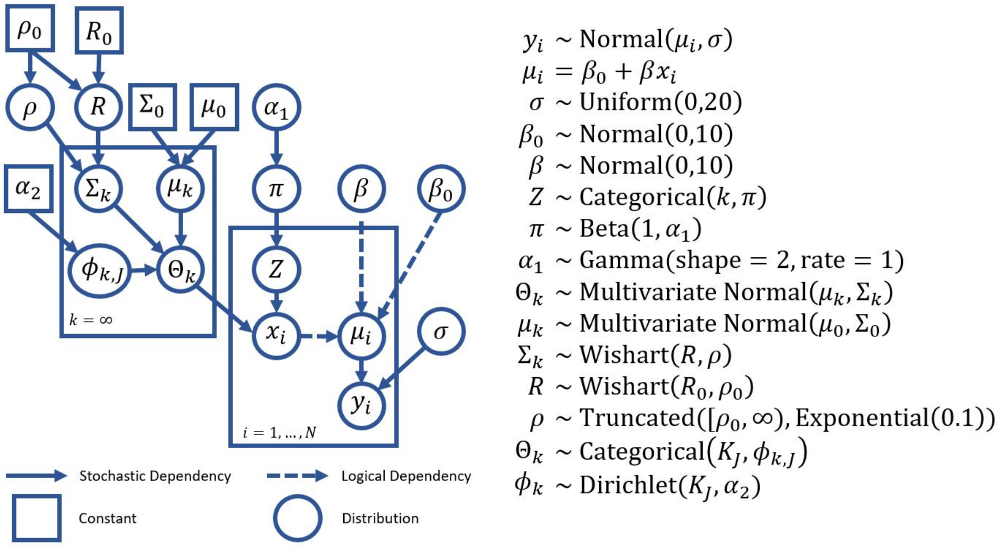


Figure S1: A Directed Acyclic Graph (DAG) of the Bayesian treatment selection model $(y_i, \boldsymbol{\mu}_i, \sigma, \boldsymbol{\beta}, \beta_{\mathbf{0}}, \mathbf{x}_i)$ augmented with a Dirichlet Process Mixture Model (DPMM). The DPMM component k assigned for each patient $\mathbf{Z}_i$, is defined by $\pi$ and $\alpha_{\mathbf{1}}$. The DPMM is given by a mixture of Gaussian and discrete variables. For Gaussian mixtures, the component specific parameters are $\boldsymbol{\mu}_k$ and $\boldsymbol{\Sigma}_k$, with other parameters as priors ($\boldsymbol{\mu}_0$,$\boldsymbol{\Sigma}_0$, $R$,$\rho$,$R_0$,$\rho_0$). For discrete mixtures, the component specific parameters is $\boldsymbol{\phi}_{kJ}$ with a flat Dirichlet prior for $K_J$ variable categories.

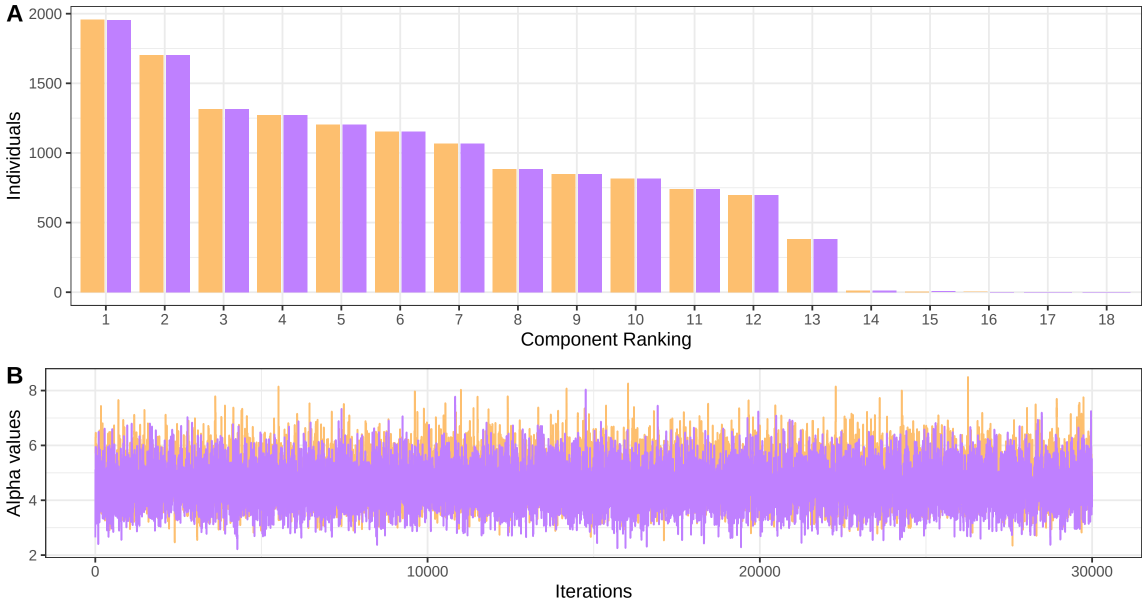


Figure S2: A: Average number of patients in components ranked by numbers of membership at each iteration after convergence. B: Trace plots of the Dirichlet process mixture model (DPMM) alpha parameter and number of components utilised for 2 chains with 30,000 iterations (20,000 iterations of burn-in removed).

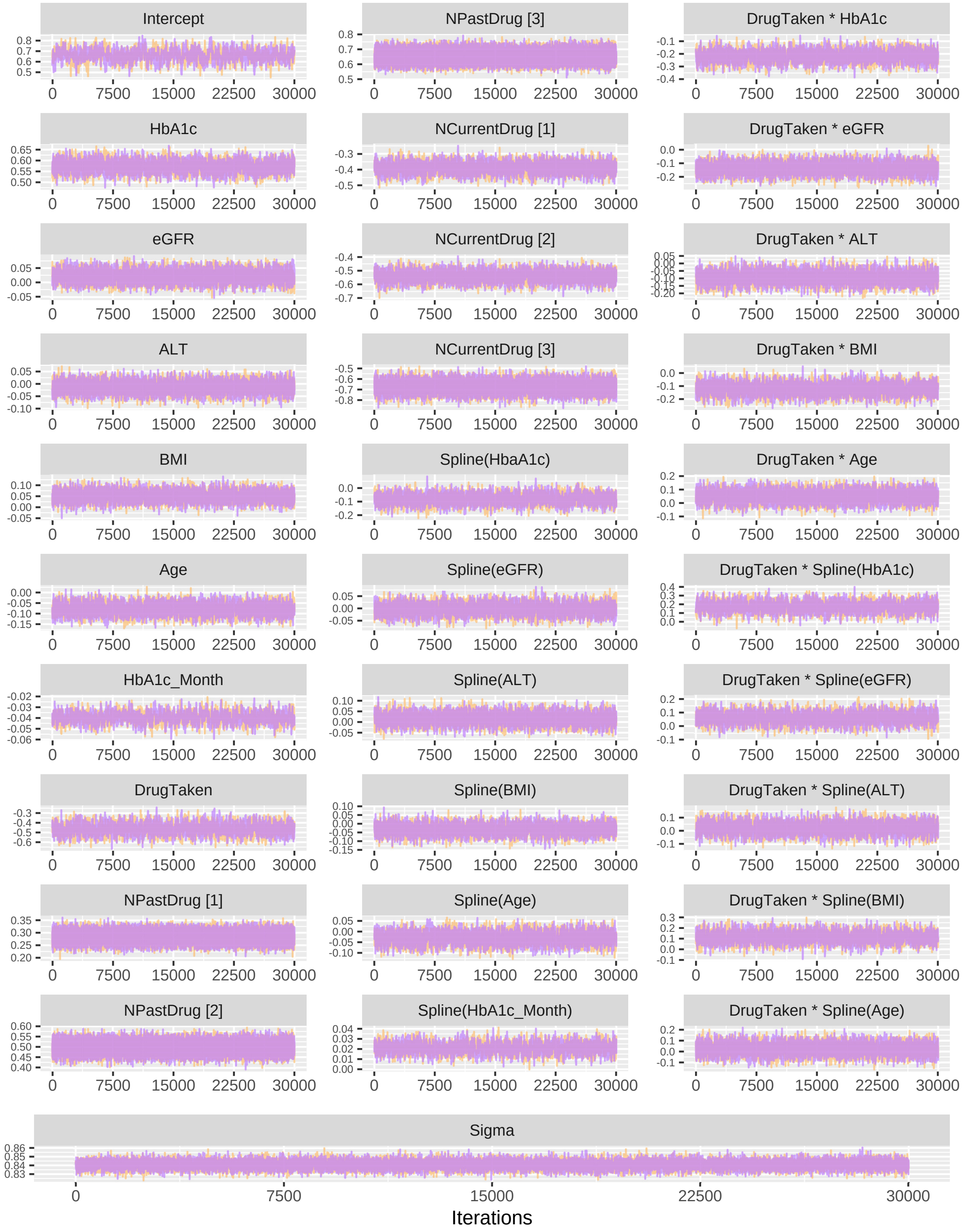


Figure S3: Trace plots of regression parameters for two chains each with 30,000 iterations (20,000 iterations of burn-in removed).

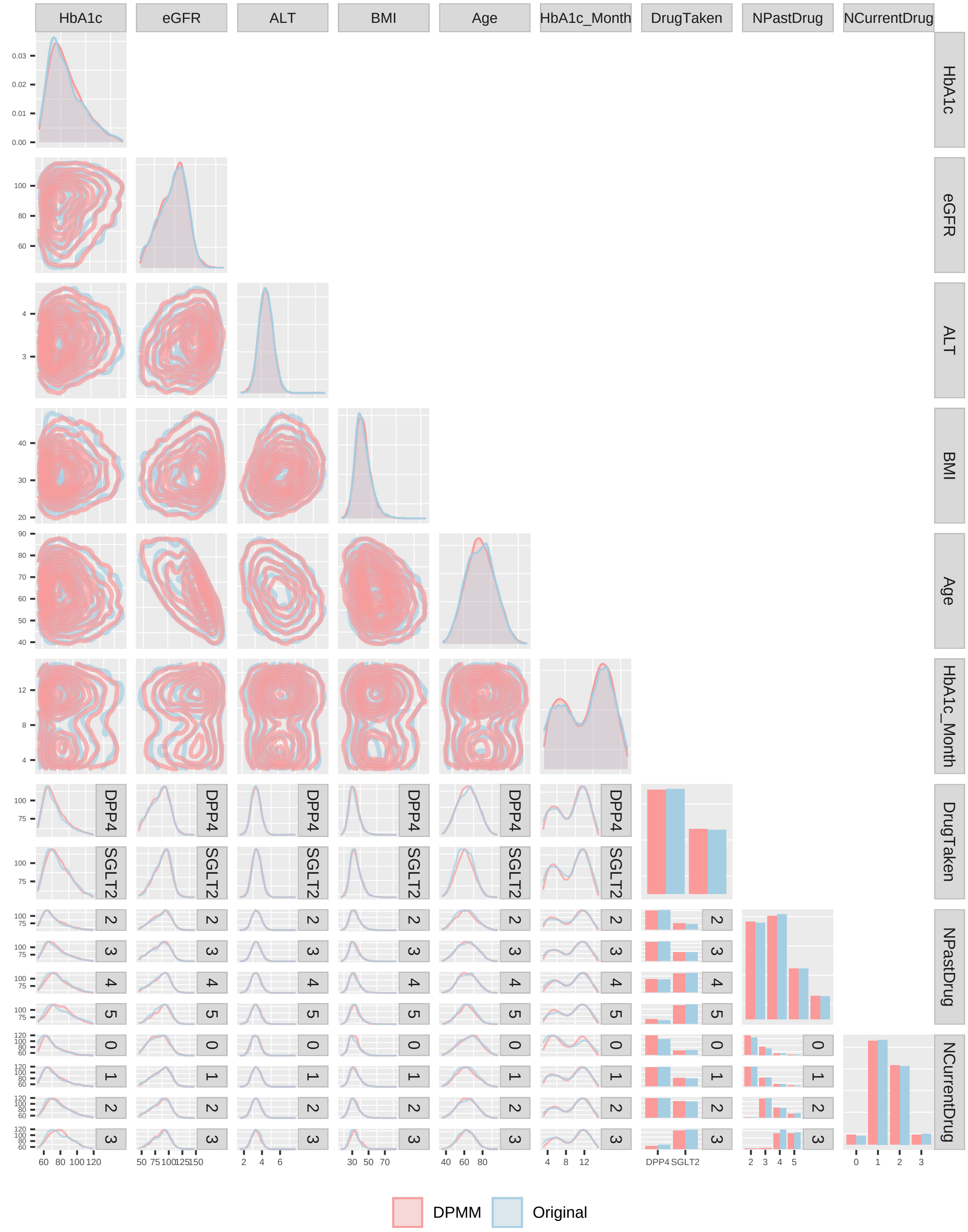


Figure S4: Generalised pairs plot of predictor variables for the development dataset against an equal number of posterior predictive samples from the Dirichlet process mixture model (DPMM).

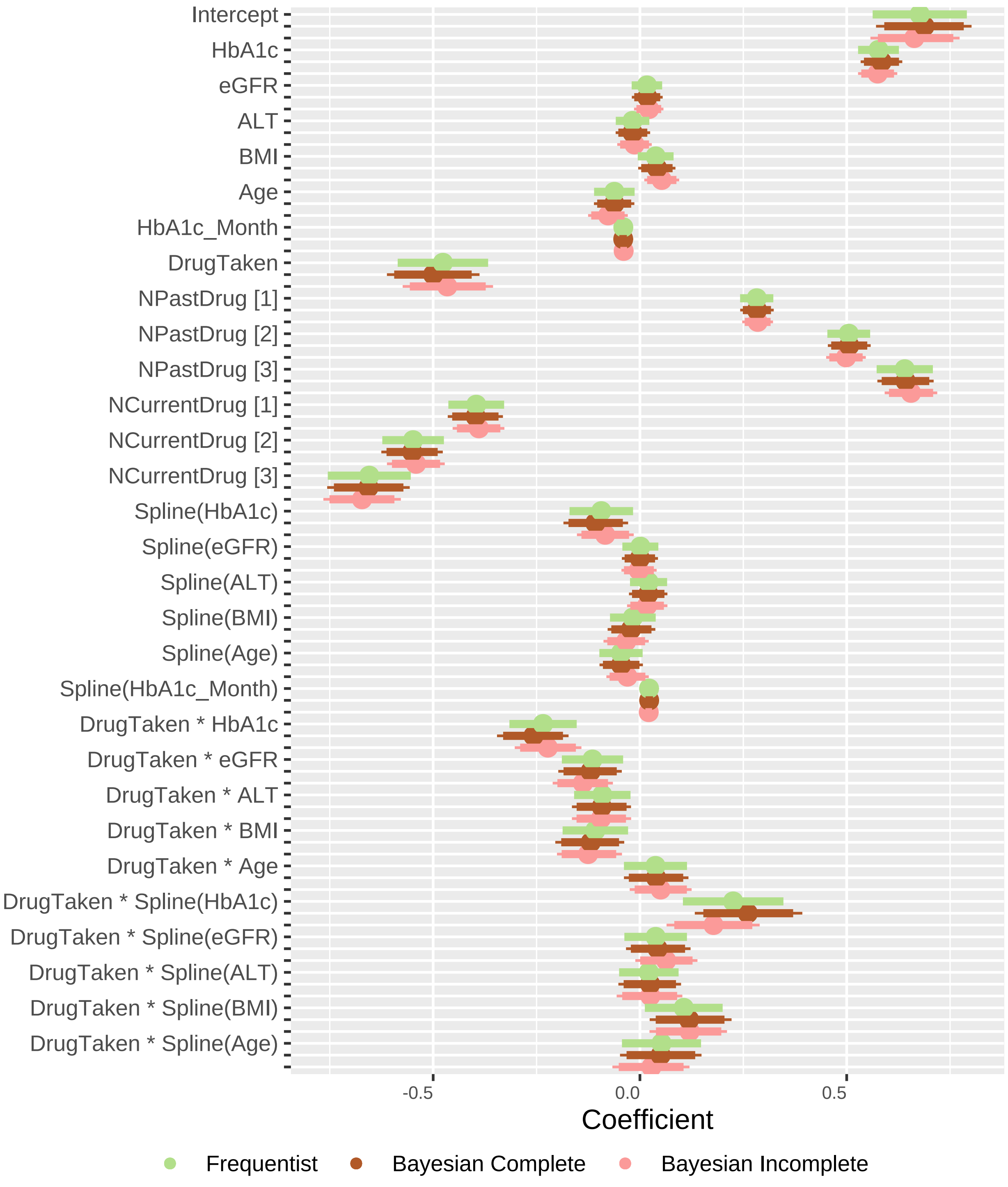


Figure S5: Caterpillar plot comparing Bayesian posterior samples fitted for the complete and incomplete datasets against the frequentist coefficient estimates. For each set of Bayesian posterior samples, the plot shows the 2.5%, 5%, 50%, 95% and 97.5% quantiles. For the frequentist estimates, the plot displays the 95% confidence interval.

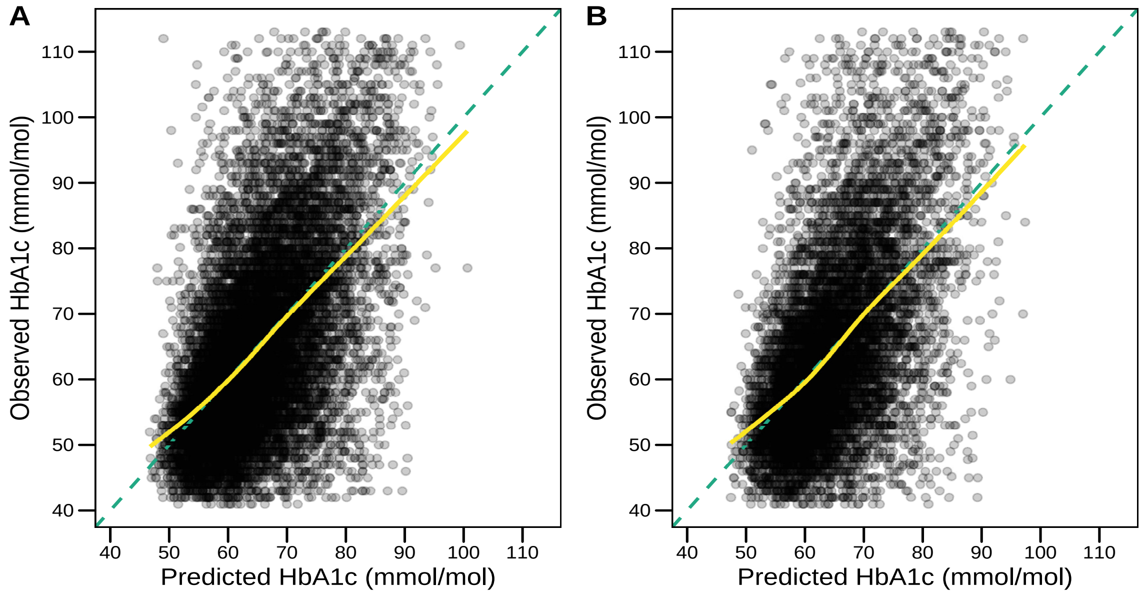


Figure S6: Predicted outcome (at 6 months) versus observed outcome for development (A) and validation (B) datasets, utilising posterior mean predictions from the fitted Bayesian treatment selection model.

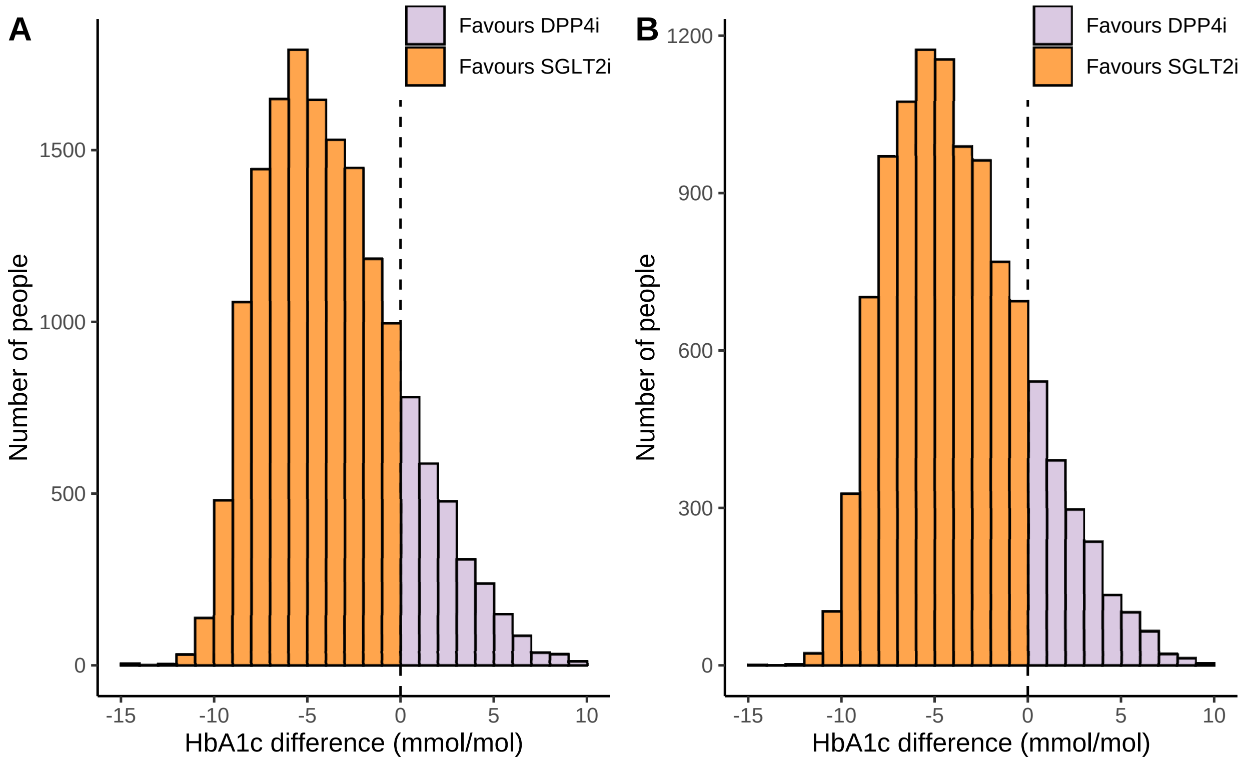


Figure S7: Comparison of individualised treatment effects for SGLT2i and DPP4i treatments in the development (A) and validation (B) datasets. A negative value corresponds to a predicted glucose-lowering treatment benefit on SGLT2i and a positive value corresponds to a predicted glucose-lowering treatment benefit on DPP4i.

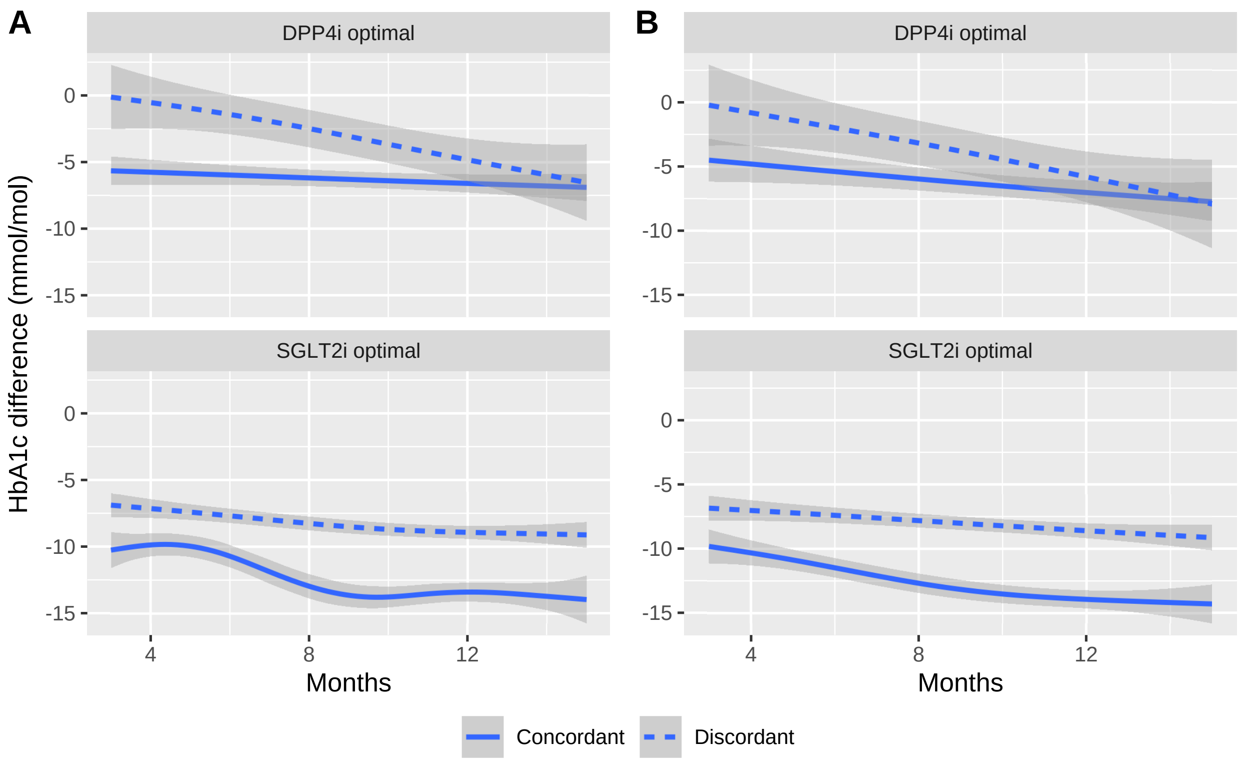


Figure S8: Therapy HbA1c reduction over time for development (A) and validation (B) datasets. The concordant subgroups experience a higher therapy benefit than the discordant subgroups.

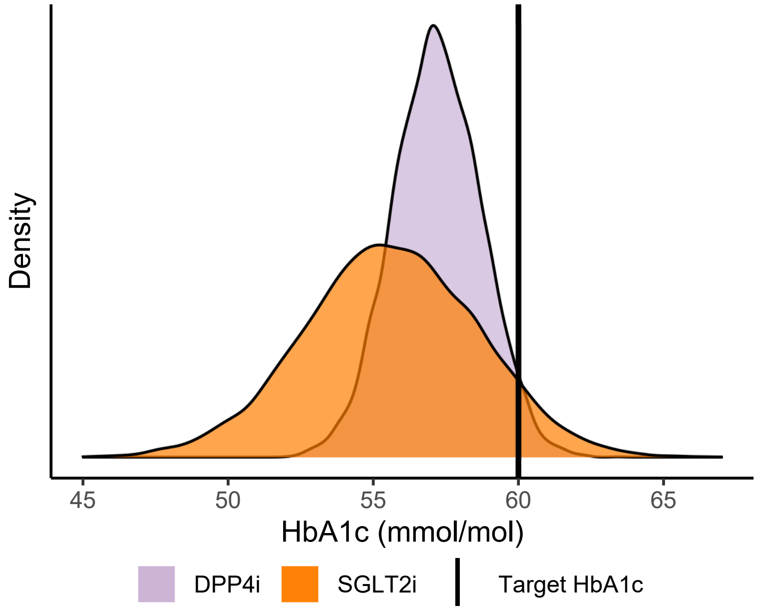


Figure S9: Therapy probability distribution at 6 month prediction for an arbitrary patient. The patient has a <60 mmol/mol target HbA1c outcome. In this situation, SGLT2i has a 7.7% chance of >60 mmol/mol treatment response, whereas DPP4i has a 3.1% chance of >60 mmol/mol. Due to the higher chance of treatment response above the target, DPP4i is the right therapy for this patient.
