## Supplementary material for "Dirichlet process mixture models to estimate outcomes for individuals with missing predictor data: application to predict optimal type 2 diabetes therapy in electronic health record data": Sample Code: Bayesian_Treatment_DPMM.pdf

This is a thorough explanation of the R code used in fitting and generating posterior predictive samples of a Bayesian Treatment Selection model augmented with a Dirichlet process mixture model (DPMM).

In this manual, we use the following libraries for: preparing the dataset for fitting, initialise values for the MCMC chains, fit the Bayesian model and making predictions.

```
library(MASS)
library(nimble)
library(tidyverse)
library(rms)
library(modelr)
library(abind)
library(coda)
library(condMVNorm)
```

The dataset included alongside this file is a synthetic version of the original dataset with only the original response and predictor variables.

```
dataset <- readRDS("sample_dataset.rds")
```

### 1. Setup dataset

The setup involves standardising all continuous variables of the model.

```
mean_values <- vector(mode = "numeric", length = 6)
sd_values <- vector(mode = "numeric", length = 6)

for (i in 1:6) {
    mean_values[i] <- mean(dataset[, i])
    sd_values[i] <- sd(dataset[, i])
}

names(mean_values) <- colnames(dataset[, c(1:6)])
names(sd_values) <- colnames(dataset[, c(1:6)])

standardisation <- list(mean_values, sd_values)
```

It is important to keep the standardisation values. These are used during prediction to back-transform the predicted values into the original scale.

```
for (i in 1:6) {
    # first we take away the mean of the variable
    dataset[, i] <- dataset[, i] - standardisation[[1]][i]
    # second we divide by the standard deviation
    dataset[, i] <- dataset[, i]/standardisation[[2]][i]
}
```

The standardisation is followed by generating all the latent variables for the categorical predictor variables.

```
y_original <- dataset %>%
    mutate(Intercept = 0) %>%
    model_matrix(Intercept ~ posthba1c_final + drugclass + npastdrug +
        ncurrentdrug + prehba1cmmol + egfr_ckdepi + prealtlog +
        prebmi + agetx + hba1cmonth) %>%
    select(-"(Intercept)")
```

It also involves calculating the knots and spline values for all continuous predictor variables. For that we use a custom function that creates new spline predictor variables and saves all knot values.

```
calc_parms <- function(dataset, nk = 3) {
    if (nk != 3)
        stop("Only works for nk = 3 currently")
    ncol <- ncol(dataset)
    parms <- matrix(0, nrow = nk, ncol = ncol)
    for (i in 1:ncol) {
        temp <- rcs(as.matrix(dataset[, i]), nk)
        parms[, i] <- attr(temp, "parms")
        dataset[, i] <- matrix(temp, ncol = 2)[, 2]
    }
    colnames(dataset) <- paste0("rcs_", colnames(dataset))
    return(list(dataset = dataset, knots = parms))
}
temp <- calc_parms(select(y_original, prehba1cmmol, egfr_ckdepi,
    prealtlog, prebmi, agetx, hba1cmonth))
y_original <- cbind(y_original, temp$dataset)
knots <- temp$knots
```

### 2. Setup Bayesian model (nimble)

First, we setup the constant values.

```
# number of categories for all categorical predictor variables
cat_vars <- dataset[,c("drugclass","npastdrug","ncurrentdrug")] %>%
  as.matrix()
ndiscdim = as.vector(apply(cat_vars, 2, function(x) length(unique(x))))

consts <- list(
  N = nrow(y_original),    # iterated through all patients
  ndim = 6,                # number of continuous variables
  ndisc = 7,               # number of categorical latent variables
  nint = 5,                # number of interactions
  ndisc_numeric = 3,       # number of starting categorical variables
  L = 5,                   # number of components in DPMM
  ndiscdim = ndiscdim      # number of categories
)
```

Second, we setup the data for the model.

```
# initial values for the DPMM
cont_vars <- dataset[, c("prehba1cmmol", "egfr_ckdepi", "prealtlog",
                         "prebmi", "agetx", "hba1cmonth")] %>%
  as.matrix()
mu0 = apply(cont_vars, 2, mean)
tau0 = apply(cont_vars, 2, range)
tau0 = solve(diag(apply(tau0, 2, diff)^2))
R0 = solve(cov(cont_vars)) / ncol(cont_vars)
rho0 = ncol(cont_vars)
delta = matrix(rep(1, consts$ndisc_numeric * max(consts$ndiscdim)),
               nrow = consts$ndisc_numeric)

data <- list(
  # response
  y = y_original[,1],
  # continuous variables
  x_cont = as.matrix(y_original[,9:14]),
  # discrete variables using latent variables for regression
  x_disc = as.matrix(y_original[,2:8]),
  # spline of continuous variables
  x_spline = as.matrix(y_original[,15:20]),
  # original discrete variables
  x_orig_disc = sapply(dataset[,c(8,9,10)], function(x) as.numeric(x)),
  # initial values for DPMM priors
  mu0 = mu0,       # initial mu values
  tau0 = tau0,     # initial tau values
  R0 = R0,         # initial R values
  rho0 = rho0,     # initial rho values
  delta = delta    # concentration parameter
)
```

Third, we setup the model.

```
# likelihood of model
code <- nimbleCode({
  ## likelihood terms
  for(i in 1:N) {
    ## DPMM for continuous
    z[i] ~ dcat(w[1:L])
    x_cont[i, ] ~ dmnorm(muL[z[i], ], prec = tauL[, , z[i]])
    ## DPMM for discrete
    for (j in 1:ndisc_numeric) {
      x_orig_disc[i, j] ~ dcat(phiL[j, 1:ndiscdim[j], z[i]])
    }
    ## Regression
    y[i] ~ dnorm(beta0 +
      inprod(beta_disc[1:ndisc], x_disc[i, 1:ndisc]) + # betas for discrete vars
      inprod(beta_cont[1:ndim], x_cont[i,1:ndim]) + # betas for continuous vars
      inprod(beta_rcs[1:ndim], x_spline[i, 1:ndim]) + # betas for spline vars
      # betas for interactions between drugclass and spline/continuous variables
      beta_int_cont[1] * x_disc[i,1] * x_cont[i,1] +
      beta_int_cont[2] * x_disc[i,1] * x_cont[i,2] +
      beta_int_cont[3] * x_disc[i,1] * x_cont[i,3] +
```

```
      beta_int_cont[4] * x_disc[i,1] * x_cont[i,4] +
      beta_int_cont[5] * x_disc[i,1] * x_cont[i,5] +
      beta_int_spline[1] * x_disc[i,1] * x_spline[i,1] +
      beta_int_spline[2] * x_disc[i,1] * x_spline[i,2] +
      beta_int_spline[3] * x_disc[i,1] * x_spline[i,3] +
      beta_int_spline[4] * x_disc[i,1] * x_spline[i,4] +
      beta_int_spline[5] * x_disc[i,1] * x_spline[i,5] , sd = sigma)
  }
  ## priors for regression
  beta0 ~ dnorm(0, sd = 2.5) # intercept
  for (k in 1:ndisc) {
    beta_disc[k] ~ dnorm(0, sd = 2.5) # betas for discrete vars
  }
  for (k in 1:ndim) {
    beta_cont[k] ~ dnorm(0, sd = 2.5) # betas for continuous vars
    beta_rcs[k] ~ dnorm(0, sd = 2.5) #  betas for spline vars

  }
  for (k in 1:nint) {
    # betas for interaction between drugclass and continuous variables
    beta_int_cont[k] ~ dnorm(0, sd = 2.5)
    # betas for interaction between drugclass and spline variables
    beta_int_spline[k] ~ dnorm(0, sd = 2.5)
  }
  sigma ~ dexp(1)
  ## priors for DPMM
  alpha ~ dgamma(shape = 2, rate = 1)
  for(i in 1:(L - 1)) {
    v[i] ~ dbeta(1, alpha)
  }
  w[1:L] <- stick_breaking(v[1:(L - 1)])
  ## hyperpriors for continuous predictors DPMM
  R1[, ] ~ dwish(R0[, ], rho0)
  rho1 ~ T(dexp(0.1), rho0, )
  for(i in 1:L) {
    muL[i, ] ~ dmnorm(mu0[], prec = tau0[, ])
    tauL[, , i] ~ dwish(R1[, ], rho1)
    ## hyperpriors for discrete predictors DPMM
    for (j in 1:ndisc_numeric) {
      phiL[j, 1:ndiscdim[j], i] ~ ddirch(delta[j, 1:ndiscdim[j]])
    }
  }
})
```

Forth, we setup the the function to generate initial values.

```
initFn <- function(ndisc, ndim, nint, y, L, N, mu0, tau0, R0,
    rho0, ndiscdim) {
    # regression parameters
    beta0 = rnorm(1, 0, 2.5)
    beta_disc = rep(rnorm(ndisc, 0, 2.5))
    beta_cont = rep(rnorm(ndim, 0, 2.5))
    beta_rcs = rep(rnorm(ndim, 0, 2.5))
    beta_int_spline = rep(rnorm(nint, 0, 2.5))
```

```
    beta_int_cont = rep(rnorm(nint, 0, 2.5))
    sigma = rexp(1, rate = 1)
    # DPMM clustering
    alpha <- rgamma(1, shape = 2, rate = 1)
    v <- rbeta(L - 1, 1, alpha)
    w <- v[1]
    for (i in 2:(L - 1)) {
        w <- c(w, v[i] * prod(1 - v[1:(i - 1)]))
    }
    w <- c(w, prod(1 - v))
    z <- rcat(N, w)
    # DPMM for discrete
    phiL <- map(1:L, function(i, ndiscdim, m, L) {
        p <- map(ndiscdim, function(n, m, L) {
            out <- numeric(m)
            out[1:n] <- nimble::rdirch(1, rep(1, n))
            out
        }, m = m, L = L)
        p <- do.call("rbind", p)
        p
    }, ndiscdim = ndiscdim, m = max(ndiscdim), L = L)
    phiL <- abind(phiL, along = 3)
    phiL <- array(phiL, dim = c(length(ndiscdim), max(ndiscdim),
        L))
    # DPMM for continuous
    R1 <- rWishart(1, rho0, R0)[, , 1]
    rho1 <- rho0 - 1
    while (rho1 < rho0) {
        rho1 <- rexp(1, 0.1)
    }
    muL <- mvrnorm(L, mu0, tau0)
    tauL <- rWishart(L, rho1, R1)
    # Pass initial values
    inits <- list(beta0 = beta0, beta_disc = beta_disc, beta_cont = beta_cont,
        beta_rcs = beta_rcs, beta_int_spline = beta_int_spline,
        beta_int_cont = beta_int_cont, sigma = sigma, alpha = alpha,
        v = v, w = w, z = z, R1 = R1, rho1 = rho1, muL = muL,
        tauL = tauL, phiL = phiL)
    inits
}
```

### 3. Run Bayesian model (nimble)

The next step is to setup, compile and configure the model with all the necessary variables that require monitoring.

```
# set up model
model <- nimbleModel(code = code, constants = consts, data = data,
    inits = initFn(consts$ndisc, consts$ndim, consts$nint, data$y,
        consts$L, consts$N, data$mu0, data$tau0, data$R0, data$rho0,
        consts$ndiscdim))
# compile the model
cmodel <- compileNimble(model)
# set monitors
```

```r
config <- configureMCMC(cmodel, monitors = c("muL", "tauL", "v",
    "alpha", "z", "beta0", "beta_disc", "beta_cont", "beta_rcs",
    "beta_int_cont", "beta_int_spline", "sigma", "phiL"), thin = 1)
# build the model
built <- buildMCMC(config)
# compile model
cbuilt <- compileNimble(built)
# run model
cbuilt$run(niter = 10000, reset = TRUE)
```

### 4. Extract posterior samples

The last step is to extract posterior samples from the model.

```r
samples <- as.matrix(cbuilt$mvSamples) %>%
    as_tibble()
```

### 5. Making predictions for an individual with missing data

First, we remove any samples that constitute burn-in.

```r
samples %>%
    select("alpha") %>%
    as.mcmc() %>%
    plot()
```

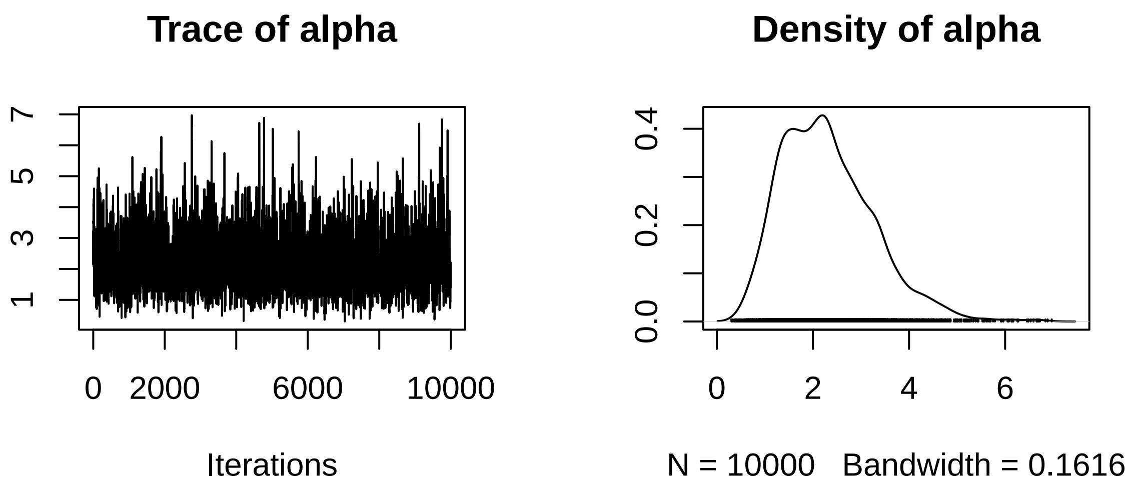


The first 5000 iterations are then removed and the remaining iterations will be used in all predictions.

```r
samples <- samples %>%
    slice(-c(1:5000))
```

The following R code is used to make predictions for missing predictor variables (`eGFR` and `NPastDrug`) and missing response (`posthba1c_final`).

Second, we prepare the posterior model samples for the DPMM parameters. That involves tidying up component samples for: weights, means, variances and categorical probabilities.

```r
# extract component weights
postW <- samples %>%
    select(starts_with("v[")) %>%
    apply(1, function(v) {
        w <- numeric(length(v) + 1)
        w[1] <- v[1]
        for (i in 2:length(v)) {
            w[i] <- v[i] * prod(1 - v[1:(i - 1)])
        }
        w[length(w)] <- prod(1 - v)
        w
    }) %>%
    t() %>%
    as_tibble() %>%
    mutate(iter = 1:n()) %>%
    gather(var, value, -iter) %>%
    mutate(var = as.numeric(gsub("V", "", var))) %>%
    arrange(iter, var) %>%
    group_by(iter) %>%
    nest() %>%
    mutate(data = map(data, "value")) %>%
    rename(w = data)
# extract component means
postMu <- samples %>%
    select(starts_with("muL")) %>%
    mutate(iter = 1:n()) %>%
    gather(var, value, -iter) %>%
    mutate(var = gsub(" |muL\\[|\\]", "", var)) %>%
    separate(var, c("component", "dim"), sep = ",") %>%
    mutate_at(vars(c("component", "dim")), as.numeric) %>%
    arrange(iter, component, dim) %>%
    select(-dim) %>%
    group_by(iter, component) %>%
    nest() %>%
    mutate(data = map(data, "value")) %>%
    group_by(iter) %>%
    nest() %>%
    mutate(data = map(data, ~{
        pluck(., "data") %>%
            abind(along = 2) %>%
            t()
    })) %>%
    rename(muL = data)
# extract component variances
postTau <- samples %>%
    select(starts_with("tauL")) %>%
    mutate(iter = 1:n()) %>%
    gather(var, value, -iter) %>%
    mutate(var = gsub(" |tauL\\[|\\]", "", var)) %>%
    separate(var, c("dim1", "dim2", "component"), sep = ",") %>%
    mutate_at(vars(c("dim1", "dim2", "component")), as.numeric) %>%
    arrange(iter, component, dim2, dim1) %>%
    select(-dim1, -dim2) %>%
```

```
    group_by(iter, component) %>%
    nest() %>%
    mutate(data = map(data, "value")) %>%
    mutate(data = map(data, ~{
        matrix(., sqrt(length(.)), sqrt(length(.)))
    })) %>%
    group_by(iter) %>%
    nest() %>%
    mutate(data = map(data, ~{
        pluck(., "data") %>%
            abind(along = 3)
    })) %>%
    rename(tauL = data)
# extract categorical probabilities
postPhi <- samples %>%
    select(starts_with("phiL")) %>%
    mutate(iter = 1:n()) %>%
    gather(var, value, -iter) %>%
    mutate(var = gsub(" |phiL\\[|\\]", "", var)) %>%
    separate(var, c("dim1", "dim2", "component"), sep = ",") %>%
    mutate_at(vars(c("dim1", "dim2", "component")), as.numeric) %>%
    arrange(iter, component, dim1, dim2) %>%
    select(-dim1, -dim2) %>%
    group_by(iter, component) %>%
    nest() %>%
    mutate(data = map(data, "value")) %>%
    mutate(data = map(data, ~{
        matrix(., nrow = 3, ncol = 4, byrow = TRUE)
    })) %>%
    group_by(iter) %>%
    nest() %>%
    mutate(data = map(data, ~{
        pluck(., "data") %>%
            abind(along = 3)
    })) %>%
    rename(phiL = data)
```

Third, we combine existing individual information with DPMM samples. At this step, we need to ensure we standardise the patients data. This is because the model was fit with standardised data.

```
dataset <- readRDS("sample_dataset.rds")
patient <- dataset[1, -1]
# missing values
patient[, c(2, 8)] <- NA
# turn categorical variables into category entry
patient[, c(7, 8, 9)] <- as.numeric(patient[, c(7, 8, 9)])
# we standardise the existing continuous variables
var_names <- colnames(patient)[c(1, 3, 4, 5)]
for (i in var_names) {
    # first we take away the mean of the variable
    patient[, i] <- patient[, i] - standardisation[[1]][i]
    # second we divide by the standard deviation
    patient[, i] <- patient[, i]/standardisation[[2]][i]
}
```

```r
# replicate patient into number of samples
patient_samples <- as.data.frame(lapply(patient, rep, nrow(samples))) %>%
    mutate(iter = 1:n()) %>%
    gather(var, value, -iter) %>%
    arrange(iter) %>%
    group_by(iter) %>%
    nest() %>%
    mutate(data = map(data, "value")) %>%
    rename(patient = data)
predictions <- patient_samples %>%
    inner_join(postW, by = "iter") %>%
    inner_join(postMu, by = "iter") %>%
    inner_join(postTau, by = "iter") %>%
    inner_join(postPhi, by = "iter")
```

Forth, we choose the component conditional on existing information.

```r
# log sum exp trick
log_sum_exp <- function(lx) {
    ## extract maximum of logged values
    mX <- max(lx)
    ## return answer
    out <- mX + log(sum(exp(lx - mX)))
    out
}

predictions <- predictions %>%
    mutate(w = pmap(list(w, muL, tauL, phiL, patient), function(w,
        mu, tau, phi, patient) {
        # which cont vars to marginalize
        continuous_patient <- patient[1:6]
        continuous_vars_given <- which(!is.na(continuous_patient))
        continuous_vars_dep <- which(is.na(continuous_patient))
        # which cat vars to iterate over
        categorical_patient <- patient[7:9]
        categorical_vars <- which(!is.na(categorical_patient))
        # marginal matrices (with var that is missing)
        marg_mean <- mu[, continuous_vars_given, drop = FALSE]
        marg_sigma <- tau[continuous_vars_given, continuous_vars_given,
            , drop = FALSE]
        lmarg <- map_dbl(1:length(w), function(i, xcont, xcat,
            mu, sigma, phi) {
            # calculate the density for continuous
            value <- mvtnorm::dmvnorm(xcont, mu[i, ], solve(sigma[,
                , i]), log = TRUE) + log(w[i])
            # calculate the density for categorical
            for (l in 1:length(categorical_vars)) {
                value <- value + log(phi[l, xcat[categorical_vars[l]],
                  i])
            }
            value

        }, xcont = patient[continuous_vars_given], xcat = patient[7:9],
            mu = marg_mean, sigma = marg_sigma, phi = phi)
```

```
        lmarg <- log_sum_exp(lmarg)

        z_given_x_l <- map_dbl(1:length(w), function(i, xcont,
            xcat, mu, sigma, phi, denom) {
            # calculate the density for continuous
            value <- mvtnorm::dmvnorm(xcont, mu[i, ], solve(sigma[,
                , i]), log = TRUE) + log(w[i]) - denom
            # calculate the density for categorical
            for (l in 1:length(categorical_vars)) {
                value <- value + log(phi[l, xcat[categorical_vars[l]],
                  i])
            }
            value
        }, xcont = patient[continuous_vars_given], xcat = patient[7:9],
            mu = marg_mean, sigma = marg_sigma, phi = phi, denom = lmarg)

        z_given_x <- exp(z_given_x_l)

        # draw cluster from adjusted probs
        draw <- which.max(rmultinom(n = 1, size = 1, prob = z_given_x))
    })) %>%
    mutate(muL = map2(w, muL, function(w, mu) {
        mu[w, ]
    })) %>%
    mutate(tauL = map2(w, tauL, function(w, tau) {
        tau[, , w]
    })) %>%
    mutate(phiL = map2(w, phiL, function(w, psi) {
        psi[, , w]
    }))
```

Fifth, we sample the missing values.

```
predictions <- predictions %>%
    mutate(value = pmap(list(patient, muL, tauL, phiL), function(patient,
        mu, tau, phi) {
        # which cont vars to marginalize
        continuous_patient <- patient[1:6]
        continuous_vars_given <- which(!is.na(continuous_patient))
        continuous_vars_dep <- which(is.na(continuous_patient))
        # which cat vars to iterate over
        categorical_patient <- patient[7:9]
        categorical_vars <- which(is.na(categorical_patient))

        preds <- NULL
        # conditional distribution for missing vars
        density <- condMVN(mu, solve(tau), dep = continuous_vars_dep,
            given = continuous_vars_given, X.given = patient[continuous_vars_given])
        # draw for missing var
        preds <- cbind(t(MASS::mvrnorm(1, density$condMean, density$condVar)))

        preds <- preds %>%
            cbind(rcat(1, phi[2, ]))
```

```
        preds <- as.data.frame(preds)
        preds
    })) %>%
    select(iter, value) %>%
    unnest(cols = value) %>%
    ungroup() %>%
    select(-iter) %>%
    set_names(c("egfr_ckdepi", "npastdrug"))
```

We can have a look at the predictions of the missing data. We must ensure we back-transform any continuous variables to the original scales.

```
# In order to return to the original scale, we do the
# reverse First we times by the standard deviation
var <- "egfr_ckdepi"
predictions[, var] <- predictions[, var] * standardisation[[2]][var]
# Second we add the mean
predictions[, var] <- predictions[, var] + standardisation[[1]][var]
```

The individual we are using in this predictions originally took 2 different drugs before the study started and had an eGFR value of 74.8.

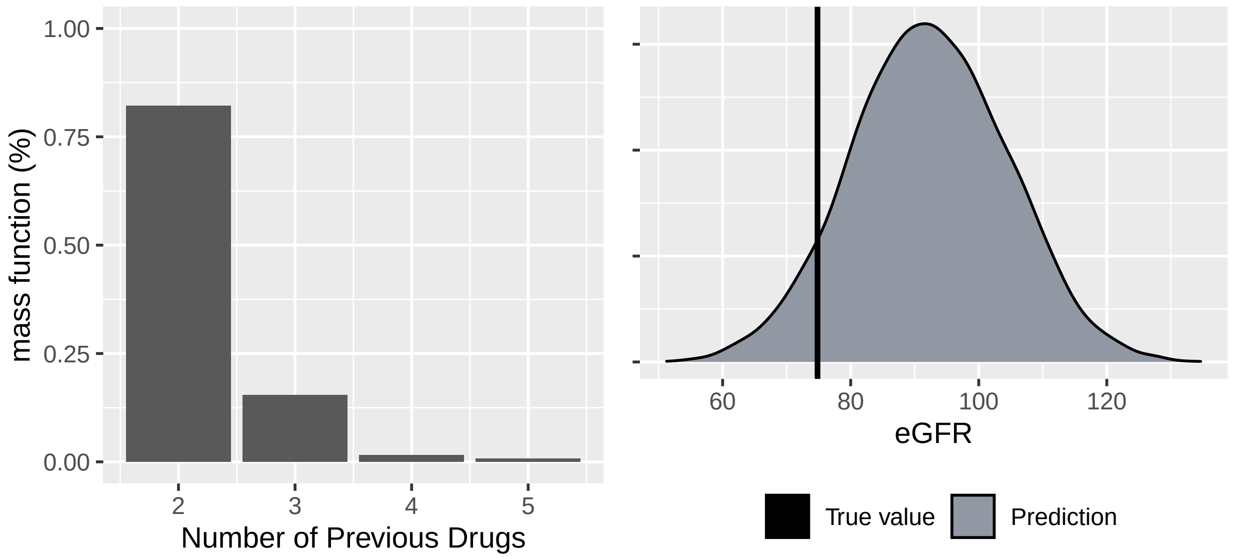

And hence we can make predictions for the outcome.

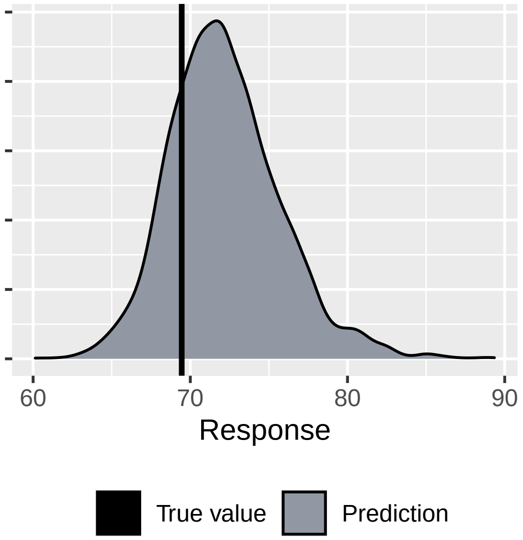
